# Identifying Contextual and Spatial Risk Factors for Post-Acute Sequelae of SARS-CoV-2 Infection: An EHR-based Cohort Study from the RECOVER Program

**DOI:** 10.1101/2022.10.13.22281010

**Authors:** Yongkang Zhang, Hui Hu, Vasilios Fokaidis, Colby Lewis V, Jie Xu, Chengxi Zang, Zhenxing Xu, Fei Wang, Michael Koropsak, Jiang Bian, Jaclyn Hall, Russell L. Rothman, Elizabeth A. Shenkman, Wei-Qi Wei, Mark G. Weiner, Thomas W. Carton, Rainu Kaushal

## Abstract

Post-acute sequelae of SARS-CoV-2 infection (PASC) affects a wide range of organ systems among a large proportion of patients with SARS-CoV-2 infection. Although studies have identified a broad set of patient-level risk factors for PASC, little is known about the contextual and spatial risk factors for PASC. Using electronic health data of patients with COVID-19 from two large clinical research networks in New York City and Florida, we identified contextual and spatial risk factors from nearly 200 environmental characteristics for 23 PASC symptoms and conditions of eight organ systems. We conducted a two-phase environment-wide association study. In Phase 1, we ran a mixed effects logistic regression with 5-digit ZIP Code tabulation area (ZCTA5) random intercepts for each PASC outcome and each contextual and spatial factor, adjusting for a comprehensive set of patient-level confounders. In Phase 2, we ran a mixed effects logistic regression for each PASC outcome including all significant (false positive discovery adjusted p-value < 0.05) contextual and spatial characteristics identified from Phase I and adjusting for confounders. We identified air toxicants (e.g., methyl methacrylate), criteria air pollutants (e.g., sulfur dioxide), particulate matter (PM_2.5_) compositions (e.g., ammonium), neighborhood deprivation, and built environment (e.g., food access) that were associated with increased risk of PASC conditions related to nervous, respiratory, blood, circulatory, endocrine, and other organ systems. Specific contextual and spatial risk factors for each PASC condition and symptom were different across New York City area and Florida. Future research is warranted to extend the analyses to other regions and examine more granular contextual and spatial characteristics to inform public health efforts to help patients recover from SARS-CoV-2 infection.

## 1. Introduction

Post-acute sequelae of SARS-CoV-2 infection (PASC) refers to ongoing, relapsing, or new symptoms occurring after the acute phase of SARS-CoV-2 infection. Approximately one in five individuals aged 18-64 and one in four individuals aged 65 or older experience potential PASC symptoms and conditions following acute SARS-CoV-2 infection (Bull-Otterson et al., 2022). Studies have identified PASC symptoms and conditions that affect multiple organ systems, including shortness of breath (Al-Aly et al., 2021; Bell et al., 2021; Taquet et al., 2021; Wang et al., 2022), fatigue (Al-Aly et al., 2021; Bell et al., 2021; Cohen et al., 2022; Shoucri et al., 2021), cognitive dysfunction (Blomberg et al., 2021; Davis et al., 2021; Taquet et al., 2021), pulmonary diseases (Cohen et al., 2022), cardiovascular diseases (Davis et al., 2021), diabetes (Cohen et al., 2022), and mental health conditions (Cohen et al., 2022; Taquet et al., 2021; Wang et al., 2022). As the number of individuals with SARS-CoV-2 infection keeps growing, understanding, treating, and preventing PASC conditions and symptoms have become a priority to help patients recover completely from SARS-CoV-2 infection. Incidence and severity of PASC symptoms and conditions vary significantly among COVID-19 patients (Groff et al., 2021; Xie et al., 2021). A critical public health objective is to identify key factors that contribute to a higher risk of PASC symptoms and conditions following SARS-CoV-2 infection. Such evidence is important to help prioritize preventions and treatment strategies and improve health equity (Sudre et al., 2021; Yoo et al., 2022). Recent studies have identified a set of patient-level risk factors for PASC among COVID-19 patients, including female sex (Bliddal et al., 2021; Sudre et al., 2021), higher body mass index (Bliddal et al., 2021; Sudre et al., 2021), older age (Carvalho-Schneider et al., 2021; Petersen et al., 2021), preexisting comorbidities (Su et al., 2022; Thompson et al., 2022), minority race/ethnicity (Halpin et al., 2021), and severity of acute SARS-CoV-2 infection (Carvalho-Schneider et al., 2021; Sudre et al., 2021). However, little is known about the environmental characteristics associated with PASC. Disadvantaged contextual and spatial characteristics, such as air pollution, social vulnerability, and poor built environment, have long been recognized as risk factors for viral respiratory infections (Diez Roux, 2001; Pica & Bouvier, 2012; Smith et al., 1999). A growing body of evidence has established strong associations between contextual and spatial risk factors (e.g., exposures to air pollutants and chemicals) and increased risk of incidence and mortality of SARS-CoV-2 infection (H. Hu et al., 2021; Weaver et al., 2022; Wu et al., 2020; Zhou et al., 2021). Recent research examined a limited set of contextual and spatial risk factors for PASC.

For example, one study examined the association between the Social Vulnerability Index (SVI) and PASC using a sample of 1,000 COVID-19 patients from a single health system and found no differences in the likelihood of PASC between patients with higher and lower levels of SVI (Yoo et al., 2022). As individuals are exposed to multiple disadvantaged contextual and spatial factors simultaneously, more research is warranted to examine the totality of the environment using COVID-19 patients from geographically diverse regions. Leveraging two large cohorts of COVID-19 patients in New York City metropolitan area and Florida, we aimed to identify contextual and spatial risk factors for a broader set of PASC symptoms and conditions associated with SARS-CoV-2 infection.

## 2. Materials and methods

### 2.1. Data Source and Setting

We conducted a retrospective cohort study using electronic health record (EHR) data from two large clinical research networks (CRNs) of PCORnet, including INSIGHT and OneFlorida+. PCORnet is a network of healthcare systems that facilitates multi-site research using EHR data. The network utilizes a common data model that fosters interoperability across participating sites. The INSIGHT CRN collects data from five academic health systems in New York City, covering a diverse patient population in the New York City Metropolitan Area (Kaushal et al., 2014). The OneFlorida+ is a partnership of 14 academic institutions and health systems across Florida, Georgia, and Alabama with longitudinal patient-level EHR data for approximately 20 million patients (Shenkman et al., 2018). Using COVID-19 patients from two regions with different social and environmental conditions helped to demonstrate the heterogeneity of contextual and spatial characteristics associated with PASC conditions.

### 2.2 Study Sample

We identified COVID-19 positive patients as those with a positive SARS-CoV-2 PCR/antigen est or COVID-19 diagnosis (U07.1, U07.2, J12.81, B34.2, B97.2, B97.21, U04, and U04.9) between March 1^st^, 2020 and October 31^st^, 2021 in both CRNs. We included COVID-19-related diagnosis codes in addition to positive laboratory test results because patients could have received a positive SARS-CoV-2 test outside CRN affiliated health systems or at home and only a diagnosis code was observed in HER data. We identified COVID-19 negative patients as those with a negative PCR/antigen test, no positive tests, and/or no COVID-19-related diagnosis codes during the same period. We defined the date of first positive or negative PCR/antigen test or COVID-19 diagnosis as the index date.

This study focused on PASC symptoms and conditions among adult patients. Patients were included if they were 20 years or older, had at least one clinical encounter 3 years to 7 days before the index date (baseline period), and had at least one encounter 31-180 days after the index date (follow-up period). This requirement was necessary to observe symptoms and conditions in the pre-test period and allow us to identify patients with incident new conditions and symptoms after SARS-CoV-2 infection.

We were also able to account for baseline demographics (e.g., age and gender) and comorbidities as confounders in the analysis. We further restricted patients to those with a 5-digit residential zip-code in EHR data. We cross-walked 5-digit zip code to 5-digit zip-code tabulation areas (ZCTA5) and only included patients from a ZCTA5 with at least ten patients. eFigures 1&2 in the appendix represented the catchment areas of our sample in New York and Florida.

### 2.3. Defining PASC

We included 23 PASC symptoms and conditions that were identified from our previous study based on existing literature, input from clinical experts, and data-driven analytics (Zang et al., 2022). A detailed description of methods of identifying these PASC symptoms and conditions was reported separately (Zang et al., 2022). These symptoms and conditions are categorized into the following eight organ systems: nervous system (encephalopathy, dementia, cognitive problems, sleep disorders, and headache), skin (hair loss and pressure ulcer of skin), respiratory system (pulmonary fibrosis, dyspnea, and acute pharyngitis), circulatory system (pulmonary embolism, thromboembolism, chest pain, and abnormal heartbeat), blood (anemia), endocrine (malnutrition, diabetes mellitus, fluid disorders, and edema), digestive system (constipation and abdominal pain), and general signs and symptoms (malaise and fatigue and joint pain). We examined contextual and spatial characteristics associated with having at least one PASC condition or symptom in each organ system as well as characteristics associated with each individual PASC condition and symptom.

### 2.4. Contextual and Spatial Characteristics

We integrated a variety of contextual and spatial measures from multiple sources to characterize patients’ exposures to their surrounding natural, built, and social environments before acute SARS-CoV-2 infection. Table 1 presents a summary of these contextual and spatial factors, along with the corresponding data sources. To account for the heterogeneous spatiotemporal scales of these factors, area- and time-weighted averages were generated to aggregate them at the ZCTA5 level. We considered a total of 259 factors covering three domains of contextual and spatial characteristics with ten categories. A complete list of factors is in the appendix (eTable 1).

**Table 1.**
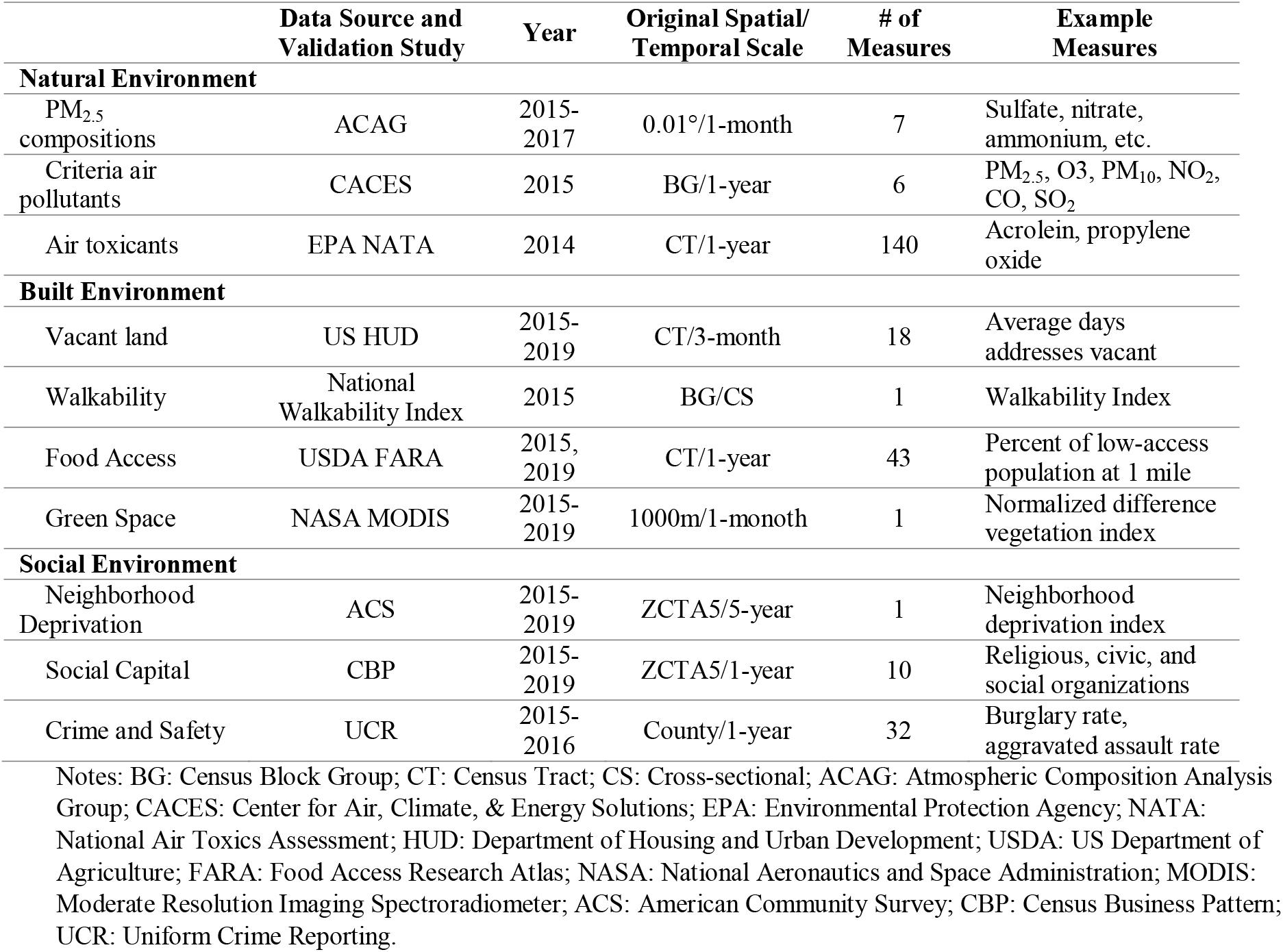
Summary of ZCTA5-level contextual and spatial characteristics.

#### 2.4.1. Natural Environment

Natural environment factors include compositions of particulate matter with diameters that are 2.5 μm and smaller (PM_2.5_ compositions), criteria air pollutants, and air toxicants. These factors could increase the risk of developing PASC by directly leading to certain conditions (e.g., respiratory diseases) or making individuals more susceptible to SARS-CoV-2 infection (e.g., exacerbate infection severity) (Weaver et al., 2022).

Data on PM_2.5_ compositions were obtained from the University of Washington at St. Louis Atmospheric Composition Analysis Group (ACAG) (van Donkelaar et al., 2019). ACAG estimated annual PM_2.5_ and its compositions at a spatial resolution of 0.01 degree in longitude and latitude. The estimates were derived using data from a chemical transport model (GEOS-Chem) and satellite observations of aerosol optical depth statistically fused by geographically-weighted models that have been extensively cross-validated (van Donkelaar et al., 2019).

We obtained criteria air pollutants, such as PM_10_ and carbon monoxide, from the Center for Air, Climate, & Energy Solutions (CACES) (S. Y. Kim et al., 2020). These measures were derived at the census block group level using data from the US Environmental Protection Agency (EPA) regulatory monitors, land use, and satellite-derived estimates of air pollution with well-validated land use regression models (S. Y. Kim et al., 2020). Finally, we obtained air toxicant measures from the National Air Toxics Assessment (NATA) conducted by EPA based on a national emissions inventory of outdoor air toxics sources (Logue et al., 2011). We used the most recent NATA data released in 2018 representing air conditions in 2014 at the census tract level. These measures represent long-term exposures rather than acute exposures to hazardous air pollutants (H. Hu et al., 2021; Petroni et al., 2020). Previous research indicates that spatial distribution of these air pollutants may have remained relatively unchanged (Chakraborty, 2021).

#### 2.4.2. Built Environment

Built environment factors, including vacant land, walkability, food access, and green space, were considered. These are important determinants to various symptoms and conditions that may be associated with SARS-CoV-2 infection. For example, better access to healthy food mitigates the risk of developing diabetes associated with SARS-CoV-2 infection (Kirby et al., 2021). Green space in neighborhood could reduce the risk of developing respiratory conditions (Tischer et al., 2017).

We obtained census-tract level vacant land measures in the period of 2015-2019 from the US Department of Housing and Urban Development (Garvin et al., 2013). We used the National Walkability Index developed by EPA, which measures walkability on a scale from 1 to 20 for each census block group, with 1 indicating the least walkable block group and 20 indicating the most walkable block group (Watson et al., 2020). Food access measures were obtained from the US Department of Agriculture (USDA)’s Food Environment Atlas (United States Department of Agriculture, 2019). We used 43 food access measures at the census-tract level of 2015 and 2019. Finally, we obtained the Normalized Difference Vegetation Index (NDVI) as a measure of green space in a neighborhood (Rhew et al., 2011). NDVI is a validated measure based on remote-sensing spectral data from NASA Moderate Resolution Imaging Spectroradiometer.

#### 2.4.3. Social Environment

We measured neighborhood deprivation, social capital, and crime and safety for neighborhood social environment (Table 1 and eTable 1). These measures represent important socioeconomic conditions that are associated with individuals’ health and various conditions.

The Neighborhood Deprivation Index (NDI) was used to characterize neighborhood socioeconomic status. NDI is a weighted average of 20 measures that represent seven domains of neighborhood deprivation, including poverty, occupation, housing, employment, education, racial composition, and residential stability. We extracted ZCTA5-level data for all 20 measures from the American Community Survey five-year estimates of 2015-2019 and derived NDI for New York, New Jersey, and Florida using an established method (Walker et al., 2020). Ten social capital measures were constructed based on the North American Industry Classification System (NACIS) codes using the 2015-2019 Census Business Pattern data at the ZCTA5-level (Rupasingha et al., 2006). Finally, we obtained county-level crime and safety measures from the Uniform Crime Reporting Program (Table 1 and eTable 1).

### 2.5. Covariates

We examined a comprehensive set of patient characteristics as potential confounders using EHR data. These included patient age (20-39 [ref.], 40-54, 55-64, 65-74, 75-84, and 85+); gender (female [ref.], male. and other/missing); race (White [ref.], Black, Asian, and other or missing); ethnicity (Hispanic [ref.], Non-Hispanic, and Missing); year-month indicators of COVID-19 positive testing (March 2020 through October 2021); baseline comorbidities; and indicators for the institutions contributing data. We used a revised list of Elixhauser comorbidities for pre-existing comorbidities, including alcohol abuse, anemia, arrythmia, asthma, cancer, chronic kidney disease, chronic pulmonary disorders, cirrhosis, coagulopathy, congestive heart failure, COPD, coronary artery disease, dementia, type 1 diabetes, type 2 diabetes, end stage renal disease on dialysis, hemiplegia, HIV, hypertension,, inflammatory bowel disorder, lupus or systemic lupus erythematosus, mental health disorders, multiple sclerosis, Parkinson’s disease, peripheral vascular disorders, pregnant, pulmonary circulation disorder, rheumatoid arthritis, seizure/epilepsy, severe obesity (BMI >= 40 kg/m2), and weight loss. Each comorbidity was identified using ICD-10-CM diagnosis codes. We also adjusted for hospitalization status for SARS-CoV-2 infection as a proxy for COVID-19 severity. Hospitalized patients were those with a hospitalization encounter in the day prior through the 16 days following the index test date whereas non-hospitalized patients were those with only an ambulatory or ED encounter in the day prior through the 16 days following the index test date.

### 2.6. Statistical Analysis

For all COVID-19 positive patients, we calculated the incidence of having at least one PASC condition in each organ system (e.g., having at least one nervous PASC condition), as well as incidence of each individual PASC condition. To calculate incidence of PASC for each organ system, we first included patients without any diagnosis of PASC conditions in that organ system during the baseline period (i.e., 3 years to 7 days before the index date). Among these patients, for each organ system we identified those with at least one diagnosis of PASC conditions during the follow-up period (i.e., 31-180 days after the index date). The incidence of PASC condition of each organ system was then calculated by dividing the number of patients in step 1 by the number of patients in step 2. Incidence of each individual PASC condition was calculated using same method by including patients without any diagnosis of a given PASC condition during the baseline period and identifying those with at least one diagnosis of that PASC condition during the follow-up period.

We derived all the 259 contextual and spatial measures for ZCTA5s in New York, New Jersey, and Florida, and merged them with EHR data of INSIGHT and OneFlorida+ CRNs. We excluded measures with five or fewer unique non-zero and non-missing values, indicating little variations in these measures across ZCTA5s in our sample. This approach led to the exclusion of 63 measures in INSIGHT sample and 55 in OneFlorida+ sample (eTable 2). The remaining 196 measures in INSIGHT and 204 in OneFlorida+ were included in our analysis. We standardized all continuous measures to account for different scales of these measures and easier interpretation.

We performed a two-phase environment-wide association study based on multiple regressions using all COVID-19 positive patients (H. Hu et al., 2021; Lin et al., 2019). We started with a data engineering process including deriving contextual and spatial measures and data linkage as mentioned above. Then in the Phase 1 analysis, we ran a single regression model for each PASC outcome (including 23 individual PASC conditions and 8 PASC groups by organ system). Each regression included one contextual or spatial factor while controlling for all covariates described above. We used mixed effects logistic regressions with a random intercept for each ZCTA5. We used the false discovery rate (FDR) adjusted p values (q values) to account for multiple testing. A contextual or spatial factor was considered significant if the q-value is < 0.05.

In Phase 2, we ran a single mixed effects logistic regression with ZCTA5 random intercepts for each PASC outcome including all the significant contextual and spatial factors identified in Phase 1, adjusting for the same set of patient level covariates. We calculated the variance inflation factor (VIF) for each PASC outcome to examine multicollinearity among all significant contextual and spatial factors and excluded factors with a VIF of 10 or higher. We identified contextual and spatial risk factors for each PASC outcome as those with a statistically significant adjusted odds ratio > 1 (P < 0.05).

Contextual and spatial characteristics could be risk factors among all patients, regardless of COVID-19 status. For example, COVID-19 negative patients could also develop respiratory conditions after long-term exposures to air pollutants. We therefore performed an additional analysis to examine the excessive risk of contextual and spatial characteristics for PASC symptoms and conditions among COVID-19 positive patients compared with negative patients. For each PASC outcome, we included both COVID-19 positive and negative patients and ran a single mixed effects logistic regression. Each regression included all the significant contextual and spatial risk factors identified from Phase 2 analysis, an indicator of COVID-19 status, an interaction term between each contextual and spatial risk factor and COVID-19 status, all other covariates, and ZCTA5 random intercepts. We identified contextual and spatial factors with excessive risk for COVID-19 positive patients if the interaction term between this factor and COVID-19 status > 1 and was statistically significant (P < 0.05). All analyses were done using R.

This study was approved by the Institutional Review Boards of Weill Cornell Medicine (21-10-95-380) and University of Florida (IRB202001831).

## 3. Results

### 3.1. Patient Characteristics

We included 65,472 COVID-19 patients from the INSIGHT CRN and 35,023 from the OneFlorida+ CRN (Table 2). OneFlorida+ had a higher proportion of patients under 65 than INSIGHT (78% vs 70%, P<0.001). Both CRNs had more female patients (60% or higher) than male patients (40% or lower). INSIGHT included a lower proportion of Black patients (18% vs 31%, P < 0.001) but a higher proportion of Hispanic patients (25% vs 17%, P < 0.001). A higher proportion of COVID-19 patients were hospitalized in OneFlorida+ than INSIGHT (25% vs 19%, P < 0.001). More patients from INSIGHT tested positive for SARS-CoV-2 in early waves of the pandemic than patients from OneFlorida+. Nearly 30% of INSIGHT patients tested positive in March to June 2020, as compared to 12% in OneFlorida+. Overall, patients from OneFlorida+ had a higher burden of baseline comorbidities compared with patients from INSIGHT (Table 2).

**Table 2.**
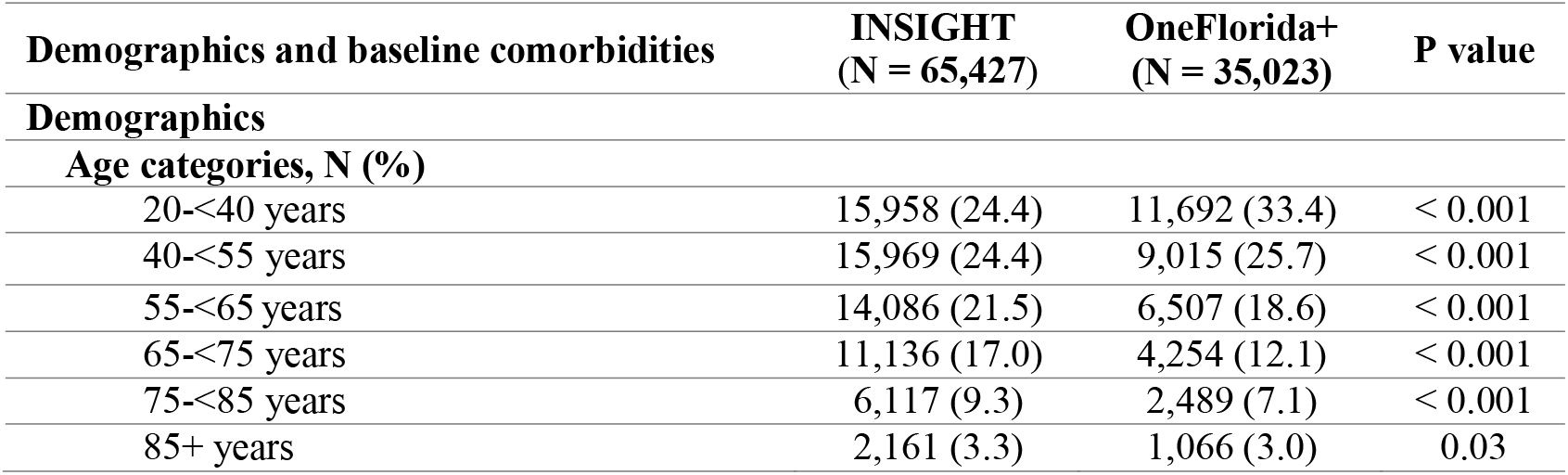
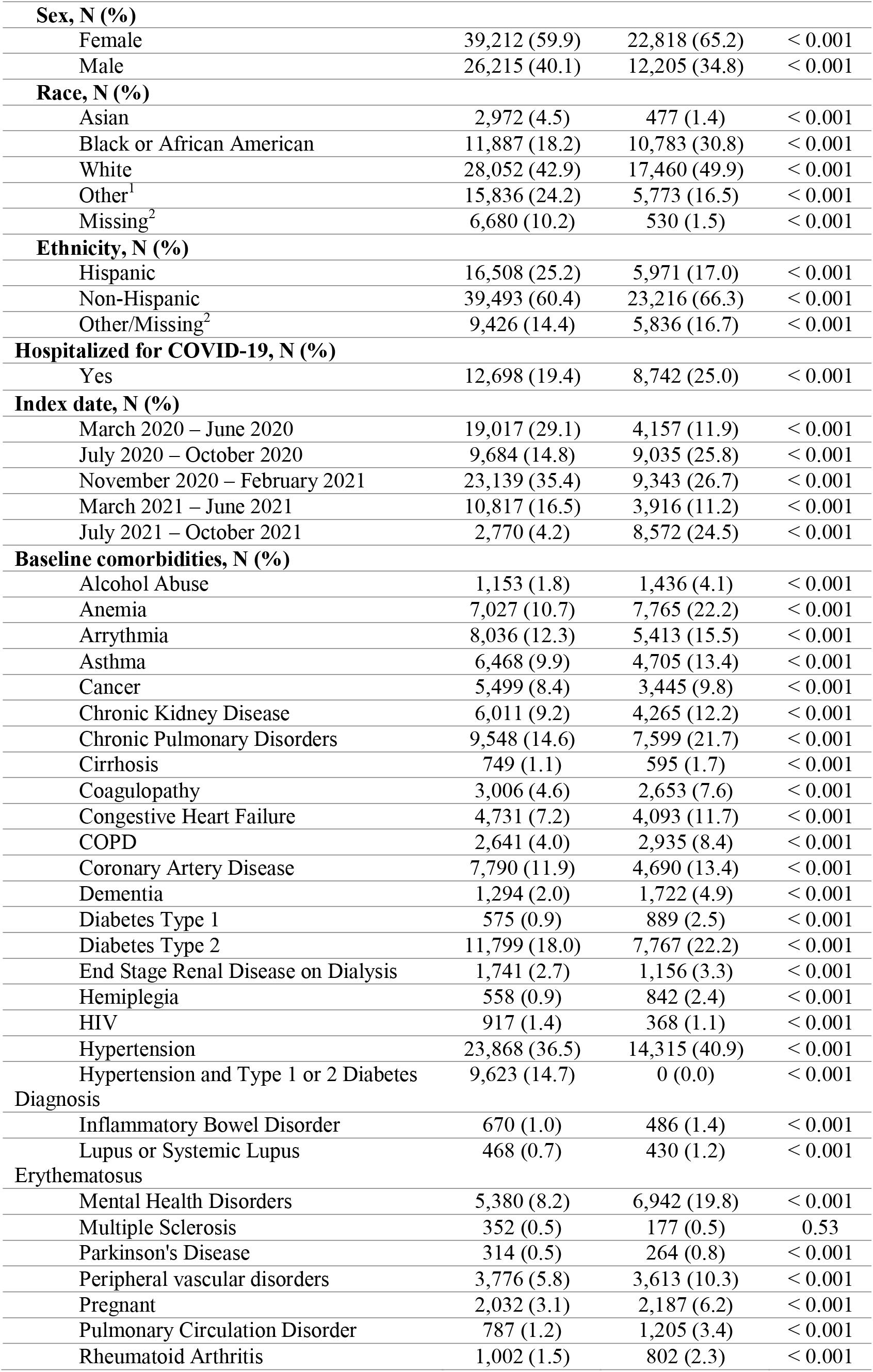
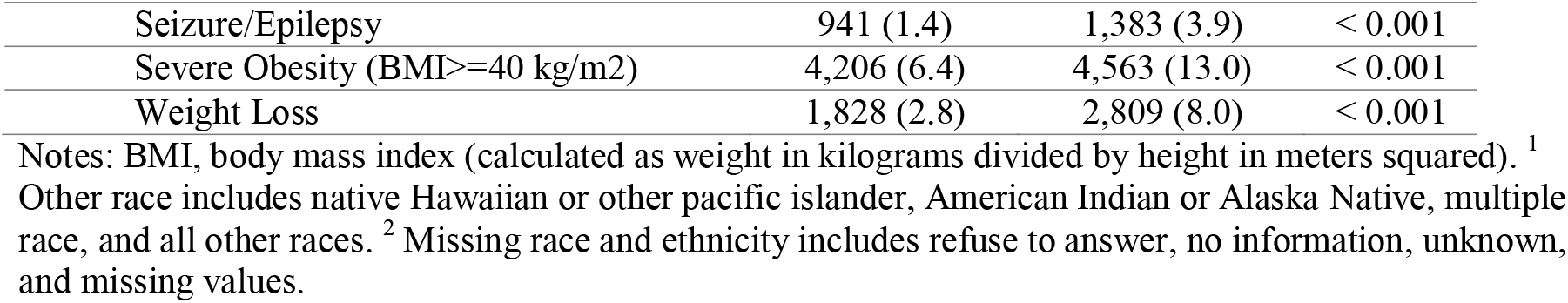
Baseline Characteristics of COVID-19 Positive Patients from INSIGHT and OneFlorida+.

### 3.2. Incidence of PASC Conditions and Symptoms

Table 3 presents incidence of PASC symptoms and conditions in both INSIGHT and OneFlorida+ cohorts among all COVID-19 positive patients. Patients from INSIGHT had higher incidence of conditions related to nervous, respiratory, circulatory, digestive, and general signs and symptoms, and lower incidence of conditions related to blood and endocrine. Incidence of conditions related to skin was similar between two CRNs. The differences in incidence of individual PASC conditions varied. Conditions with higher relative differences between INSIGHT and OneFlorida+ included fluid and electrolyte disorders (0.5% vs 4.3%, P<0.001), hair loss (1.2% vs 0.6%, P<0.001), pressure ulcer of skin (0.6% vs 1.1%, P<0.001), and acute pharyngitis (1.3% vs 1.9%, P<0.001).

**Table 3.** Incidence of New Conditions and Symptoms among COVID-19 Patients from INSIGHT and OneFlorida+.

| PASC conditions and symptoms | INSIGHT (%) | OneFlorida+ (%) | P value |
| --- | --- | --- | --- |
| **Nervous** |  |  |  |
| Encephalopathy | 1.6 | 2.1 | < 0.001 |
| Dementia | 0.8 | 1.1 | < 0.001 |
| Cognitive problems | 3.5 | 3.4 | 0.49 |
| Sleep disorders | 3.5 | 3.0 | < 0.001 |
| Headache | 3.3 | 3.8 | < 0.001 |
| Any nervous condition | 9.6 | 8.1 | < 0.001 |
| **Skin** |  |  |  |
| Hair loss | 1.2 | 0.6 | < 0.001 |
| Pressure ulcer of skin | 0.6 | 1.1 | < 0.001 |
| Any skin conditions | 1.8 | 1.7 | 0.13 |
| **Respiratory** |  |  |  |
| Pulmonary fibrosis | 2.6 | 2.5 | 0.17 |
| Dyspnea | 11.4 | 9.1 | < 0.001 |
| Acute pharyngitis | 1.3 | 1.9 | < 0.001 |
| Any respiratory condition | 13.1 | 10.4 | < 0.001 |
| **Circulatory** |  |  |  |
| Pulmonary embolism | 0.7 | 1.0 | < 0.001 |
| Thromboembolism | 1.2 | 1.3 | 0.16 |
| Chest pain | 5.6 | 5.1 | 0.005 |
| Abnormal heartbeat | 5.0 | 4.6 | 0.02 |
| Any circulatory condition | 8.9 | 8.4 | < 0.001 |
| **Blood** |  |  |  |
| Anemia | 3.9 | 4.7 | < 0.001 |
| **Endocrine** |  |  |  |
| Malnutrition | 1.3 | 1.9 | < 0.001 |
| Diabetes mellitus | 3.0 | 2.5 | < 0.001 |
| Fluid disorders | 0.5 | 4.3 | < 0.001 |
| Edema | 6.1 | 7.6 | < 0.001 |
| Any endocrine condition | 8.8 | 9.1 | 0.25 |
| **Digestive** |  |  |  |
| Other constipation | 3.3 | 2.8 | < 0.001 |
| Abdominal pain | 7.8 | 8.2 | 0.07 |
| Any digestive condition | 9.3 | 8.7 | 0.008 |
| **General signs and symptoms** |  |  |  |
| Malaise and fatigue | 4.6 | 5.0 | 0.03 |
| Joint pain | 9.7 | 7.4 | < 0.001 |
| Any general signs and symptoms | 13.0 | 9.5 | < 0.001 |

### 3.3. Contextual and Spatial Risk Factors for PASC Conditions and Symptoms

Figures 1 presents contextual and spatial factors that were significantly (q < 0.05) associated with having at least one PASC condition or symptom in each organ system from the Phase 1 analysis using COVID-19 patients from INSIGHT. One air toxicant factor was associated with respiratory PASC. A large group of air toxicant factors had significant associations with PASC related to endocrine, nervous, skin, and general signs and symptoms. In addition, food access had statistically significant associations with PASC related to endocrine, nervous, skin, and general signs and symptoms. Food access, green space, neighborhood deprivation, social capital, and vacant land were associated with PASC conditions and symptoms of endocrine, nervous, skin, and general signs and symptoms.

**Figure 1.**
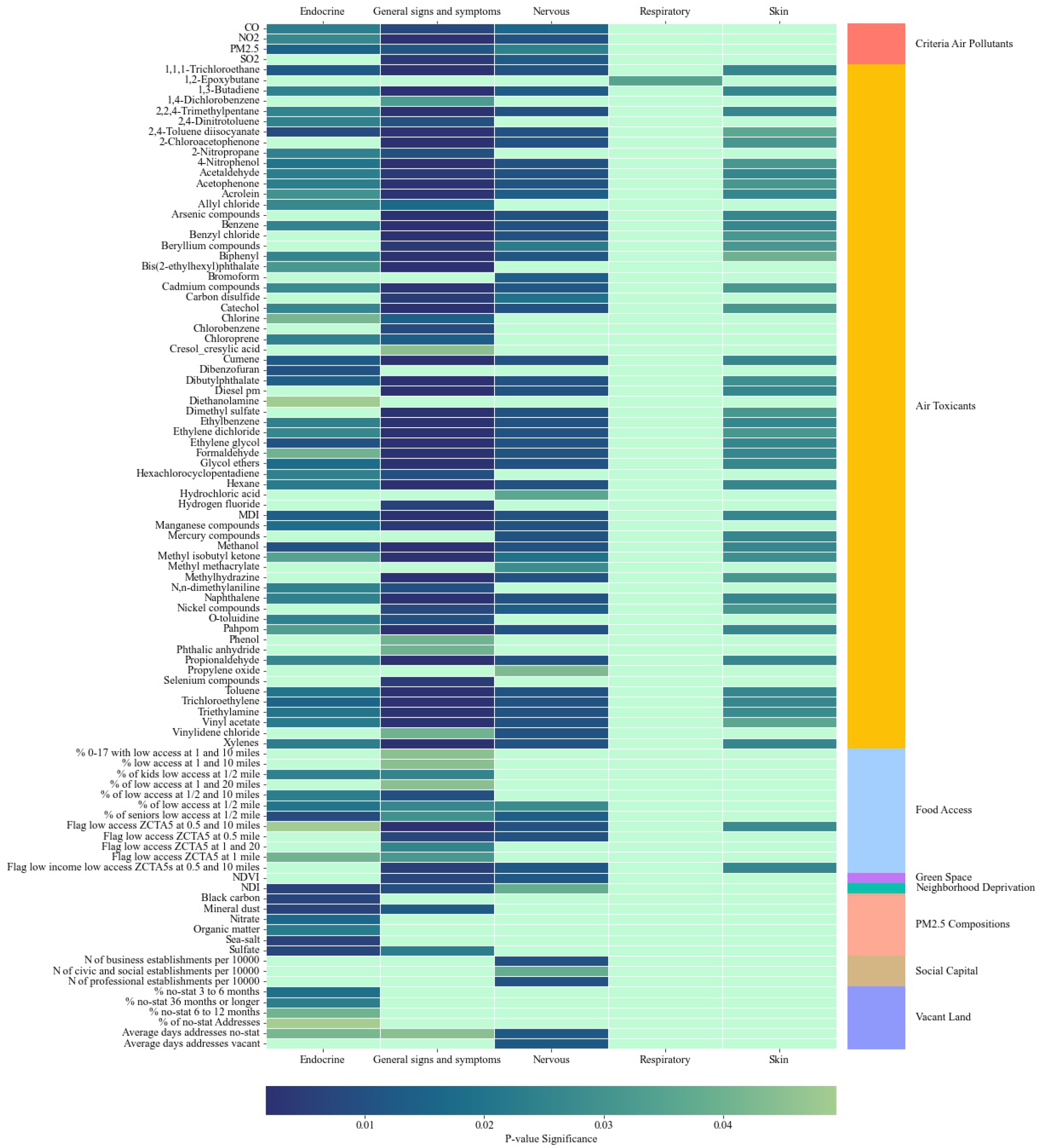
Significant Contextual and Spatial Factors Associated with PASC Groups in Phase 1 Analysis Using INSIGHT Sample. Notes: Figure represent significant neighborhood and environmental characteristics identified from mixed effects logistic regressions where a PASC condition is the outcome and each neighborhood and environmental characteristic is the key independent variable. All regressions controlled for patient-level covariates. A neighborhood or environmental characteristic is considered significant if the false discovery rate adjusted p value is < 0.05.

Figures 2 presents Phase 1 results using COVID-19 patients from OneFlorida+. Blood and skin PASC were each associated with a single air toxicant factor. Similar with INSIGHT, a large set of criteria air pollutant and air toxicant characteristics were associated with endocrine and nervous PASC. Many criteria air pollutants and air toxicants were associated with circulatory, digestive, and respiratory PASC. A smaller set of built and social environment characteristics were associated with of circulatory, digestive, endocrine, and respiratory PASC conditions and symptoms among OneFlorida+ patients.

**Figure 2.**
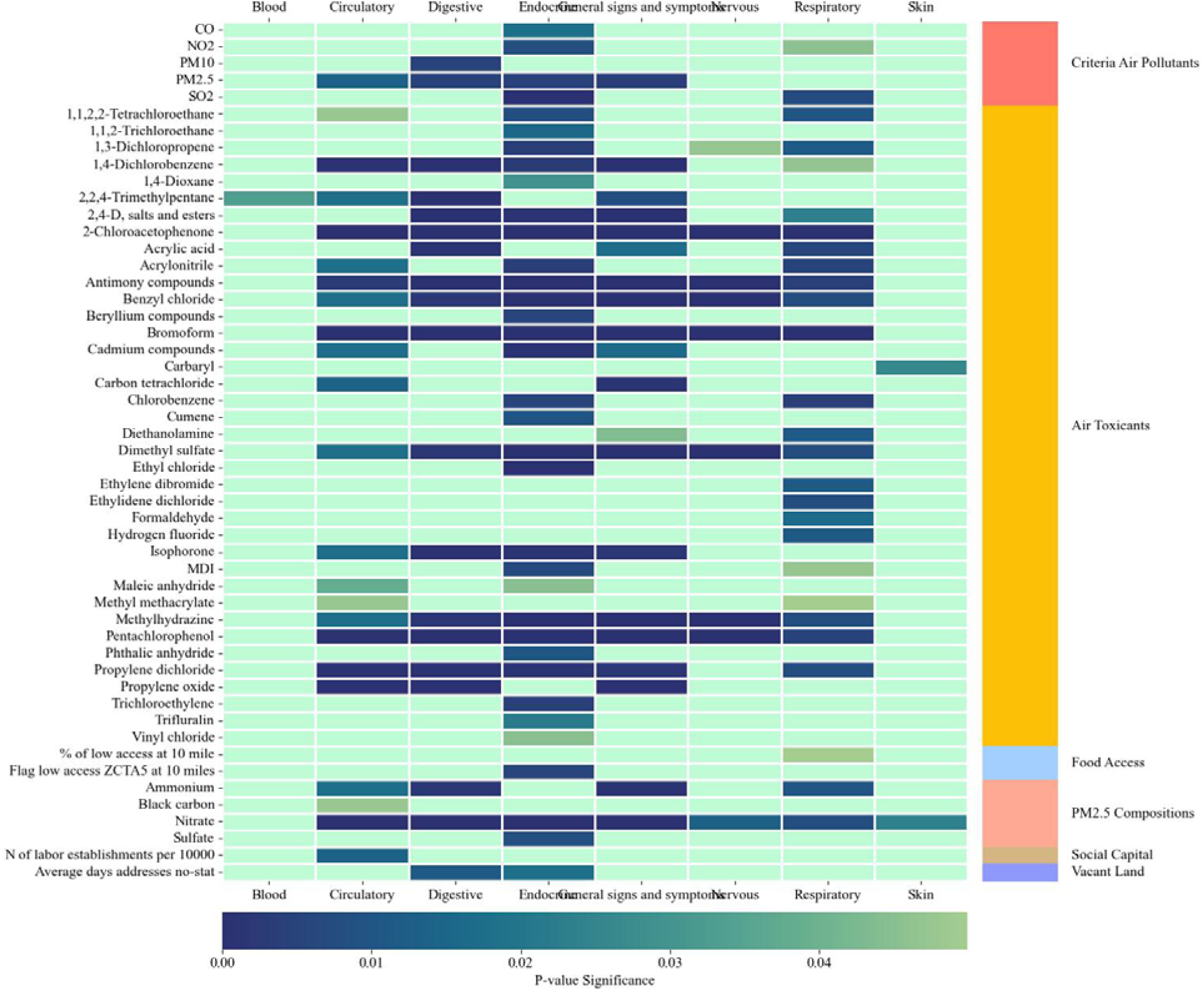
Significant Contextual and Spatial Factors Associated with PASC Groups in Phase 1 Analysis Using OneFlorida+ Sample. Notes: Figure represent significant neighborhood and environmental characteristics identified from mixed effects logistic regressions where a PASC condition is the outcome and each neighborhood and environmental characteristic is the key independent variable. All regressions controlled for patient-level covariates. A neighborhood and environmental characteristic is considered significant if the false discovery rate adjusted p value is < 0.05.

Figures 3&4 present significant contextual and spatial risk factors from Phase 2 analysis. Among COVID-19 patients from INSIGHT, we found that a higher level of air toxicants was associated with PASC conditions related to nervous, skin, and respiratory. Higher levels of methyl methacrylate in the air were associated with an increased risk of developing at least one nervous PASC condition (adjusted odds ratio [aOR]: 1.04, 95% confidence interval [CI]: 1.01-1.06). Higher neighborhood deprivation was associated with an increased risk of developing PASC of endocrine (aOR: 1.08, 95% CI: 1.02-1.15).

**Figure 3.**
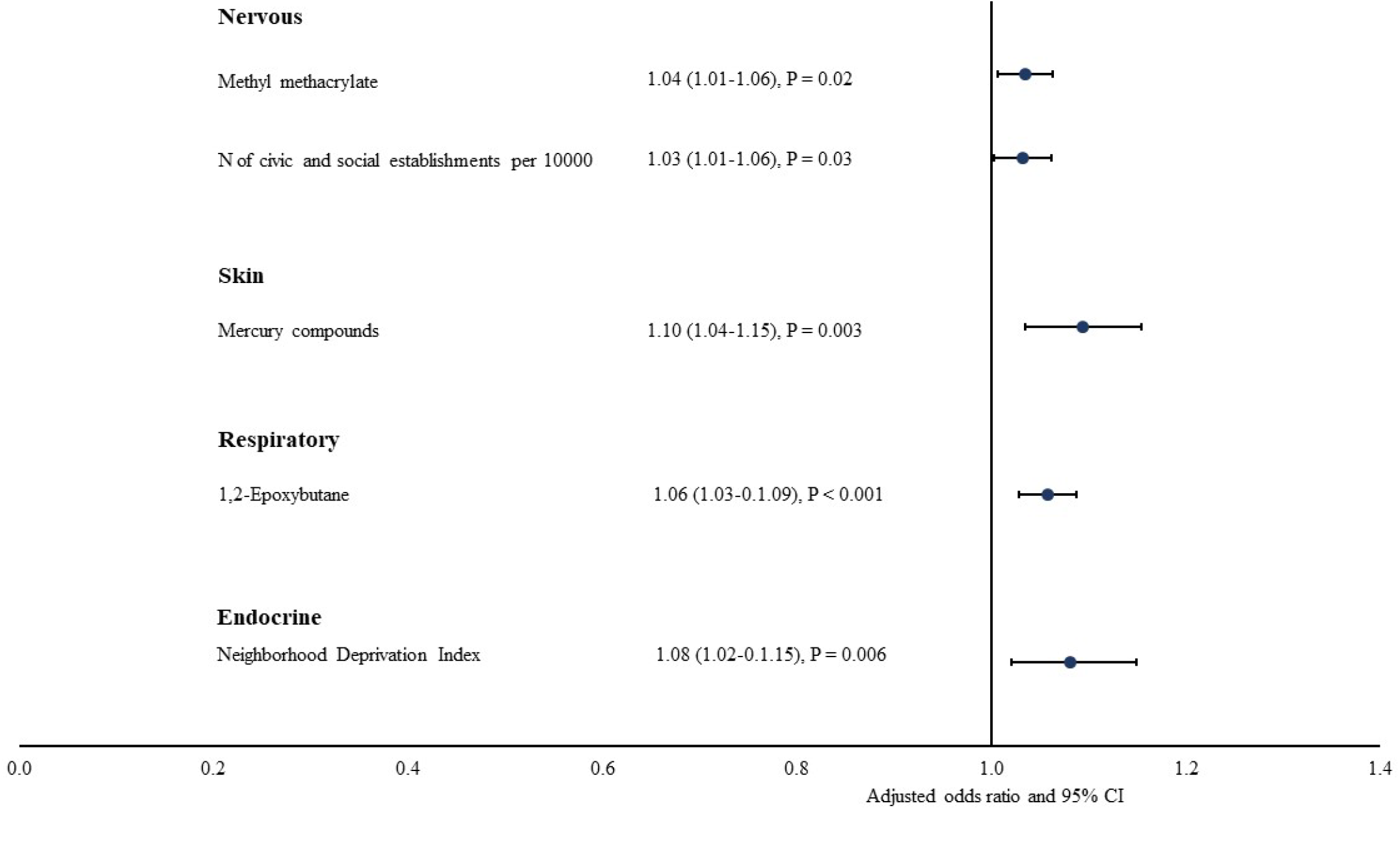
Contextual and Spatial Risk Factors for PASC Conditions by Organ System using INSIGHT Sample. Notes: NDI: Neighborhood Deprivation Index. ORs were estimated from mixed effects logistic regressions with ZCTA5 random intercept. Each regression includes all significant neighborhood and environmental characteristics identified from phase 1 analysis for each PASC outcome, controlling for all patient-level covariates.

**Figure 4.**
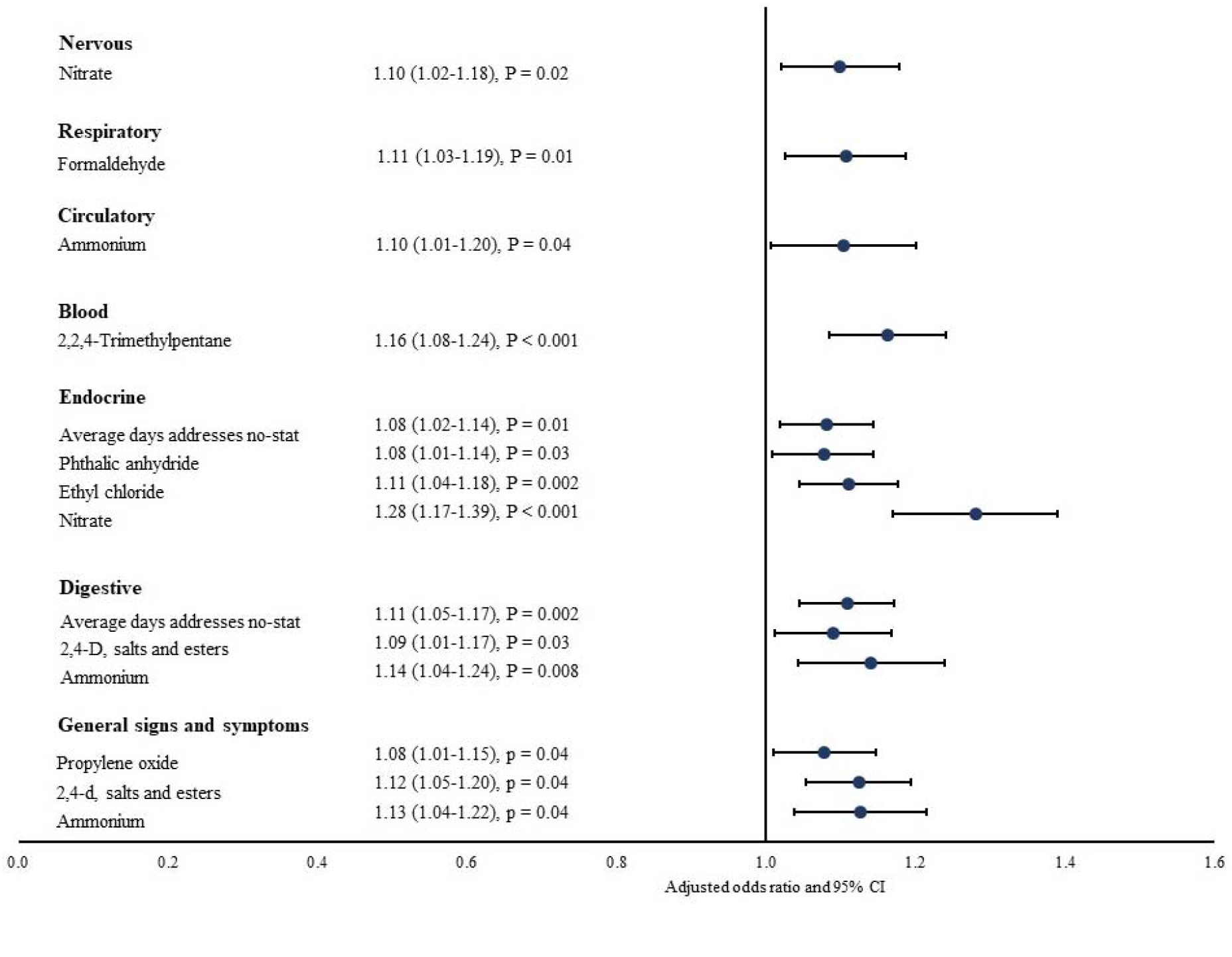
Contextual and Spatial Risk Factors for PASC Conditions by Organ System using OneFlorida+Sample. Notes: NDI: Neighborhood Deprivation Index. ORs were estimated from mixed effects logistic regressions with ZCTA5 random intercept. Each regression includes all significant neighborhood and environmental characteristics identified from phase 1 analysis for each PASC outcome, controlling for all patient-level covariates.

Using COVID-19 patients from OneFlorida+, we found that PM_2.5_ compositions were associated with increased risk of developing PASC conditions of nervous, circulatory, endocrine, digestive, and general signs. For example, a higher level of ammonium was associated with an increased risk of developing circulatory PASC (aOR: 1.10, 95% CI: 1.01-1.20). Many air toxicants were associated with an increased risk of PASC conditions affecting many organ systems, including nervous, skin, respiratory, blood, endocrine, digestive, and general signs. Average days addresses no-stat was associated with an increased risk of developing endocrine and digestive PASC.

### 3.4. Contextual and Spatial Risk Factors for Individual PASC Symptoms and Conditions

We also identified contextual and spatial risk factors for each individual PASC condition using the same analytic strategies (eFigures 3-6). Using COVID-19 patients from INSIGHT, we found that higher level of neighborhood deprivation was associated with increased risk of headache (aOR: 1.09, 95% CI: 1.02-1.16), chest pain (aOR: 1.07, 95% CI: 1.01-1.07), diabetes (aOR: 1.10, 95% CI: 1.02-1.20), and joint pain (aOR: 1.06, 95% CI: 1.01-1.11). A set of air toxicants were associated with an increased risk for encephalopathy, cognitive problems, chest pain, and other PASC conditions. Using COVID-19 from OneFlorida+ identified a broader set of air toxicants and PM _2.5_ compositions associated with an increased risk for multiple PASC conditions. For example, nitrate and ammonium were associated with an increased risk of headache, dyspnea, acute pharyngitis, and abdominal pain. Certain built environment and food access factors were also associated with certain PASC conditions in OneFlorida+ sample. Low food access of housing unit without vehicle access was associated with increased risk of fatigue (aOR: 1.08, 95% CI: 1.02-1.14).

### 3.5. Excessive Risk of Contextual and Spatial Characteristics for PASC Symptoms and Conditions

Analyses including COVID-19 negative patients and interaction terms between contextual and spatial risk factors and COVID-19 status identified several characteristics with excessive risk for PASC among COVID-19 positive patients relative to negative patients (odds ratio of the interaction term > 1 and P < 0.05). For example, we found that 1,2-epoxybutane was associated with excessive risk for respiratory PASC among COVID-19 positive patients compared with negative patients (aOR: 1.07, P < 0.001). For individual PASC symptoms and conditions, ethylene dibromide was associated with excessive risk for encephalopathy among COVID-19 positive patients compared with negative patients (aOR: 1.13, P < 0.001). Full results of these analyses are available in the appendix (eTables 3-6).

## 4. Discussions

To our knowledge, this is the first study examining contextual and spatial risk factors for a comprehensive set of PASC symptoms and conditions. Using large and diverse COVID-19 patient samples from two CRNs, we identified ZCTA5-level risk factors from nearly 200 variables for 23 PASC conditions of eight organ systems. Risk factors for PASC symptoms and conditions were primarily concentrated on air toxicants, overall neighborhood deprivation, and PM_2.5_ compositions (e.g., nitrate and ammonium). A few built environment characteristics, such as food access, were also associated with PASC symptoms and conditions. Our findings indicated significant heterogeneity in contextual and spatial risk factors for PASC between the New York City area and Florida.

Disadvantaged contextual and spatial characteristics can increase the risk for PASC through multiple direct and indirect pathways. Long-term exposure to air pollution can directly cause various symptoms and conditions of central nervous system, respiratory, endocrine, and other organ systems. The association between air pollution and respiratory conditions has been well established. PM_2.5_ is associated with increased risk of incident asthma, COPD, and other respiratory diseases (Tiotiu et al., 2020; Z. Zhang et al., 2021). Growing numbers of studies also demonstrate associations between air pollution and nervous conditions. Air pollution is associated with metabolic abnormalities and oxidative stress in the brain (H. Kim et al., 2020; Thomson, 2019). Air pollution-induced dysfunction of the insulin signaling system can reduce cognitive function and increase the risk of dementia (H. Kim et al., 2020; Paul et al., 2020). People living in neighborhoods of greater deprivation often have fewer financial resources, lower health literacy, and higher food insecurity, leading to the development of diabetes and other conditions (M. D. Hu et al., 2021; Kurani et al., 2021). Previous studies found that COVID-19 patients are disproportionately from areas with disadvantaged neighborhood conditions (Y. Zhang et al., 2021).

Addressing neighborhood and environmental vulnerability is important to help patients recover from SARS-CoV-2 infection. Compared with the robust evidence on direct health effects of contextual and spatial risk factors, the interactions between these characteristics and SARS-CoV-2 infection are understudied and may be of great importance to address. Early evidence indicated that air pollution can modify individuals’ susceptibility to SARS-CoV-2 infection and disease severity (Chen et al., 2022; Pica & Bouvier, 2012; Weaver et al., 2022). This may be mediated by upregulation of proteins critical to viral entry and by immune system suppression from oxidative stress, epithelial damage, and pulmonary inflammation (van der Valk & In ‘t Veen, 2021; Weaver et al., 2022). Studies found that exposure to particulate matter can increase the expression of angiotensin-converting enzyme 2 (ACE2) and other proteins critical to SARS-CoV-2 entry into host cells (Hoffmann et al., 2020; Sagawa et al., 2021). Upregulation of proteins necessary for viral entry may lead to higher viral load and elevate the risk of severe COVID-19.

Immunological impairment prior to COVID-19 infection, induced by long-term exposure to PM, NO_2_, and other air pollutants, may also increase the risk of COVID-19 infection and/or its severity (Weaver et al., 2022). Severe COVID-19 is associated with high inflammation and elevated levels of inflammatory cytokines, both are important pathophysiologic factors for PASC symptoms and conditions (Mehandru & Merad, 2022; Nalbandian et al., 2021). Our analyses provided important evidence to this question. Results indicated that certain contextual and spatial characteristics, particularly air toxicants, were associated with excessive risk for PASC symptoms and conditions among COVID-19 positive patients compare with negative patients.

We found considerable heterogeneity of contextual and spatial risk factors for PASC between New York City and Florida. This could be due to different neighborhood and environmental characteristics between two regions. For example, food access may be easier for patients in New York area because of the public transportation and urbanity compared with Florida. Therefore, low food access among households without vehicle access was found to be a risk factor for PASC among patients from Florida but not in New York area. A recent study also reported different levels of O_3_ pollution between New York and Florida and found different associations between O_3_ pollution and COVID-19 infection (Razzaq et al., 2020). The differential burden of preexisting comorbidities among patients in Florida may also account for the heterogeneous findings. Patients with a higher burden of pre-existing chronic conditions may be more susceptible to air pollution induced adverse health effects and therefore are at a higher risk for PASC (To et al., 2015). Other potential explanations may include variations in vaccination rate, healthcare utilization pattern, and differing courses of pandemic in these two regions. More research is needed to extend the analyses to other regions and understand reasons for heterogeneity in contextual and spatial risk factors for PASC.

This study has several major strengths. We were able to account for simultaneous exposure to multifaceted disadvantaged environmental risk factors by examining a very comprehensive set of contextual and spatial characteristics. Lack of detailed patient level data has been considered a major limitation in previous studies examining environmental risk factors and COVID-19 related outcomes (Weaver et al., 2022). Compared with previous ecologic studies relying on data aggregated at the county level, we were able to adjust for detailed patient level characteristics (e.g., demographics and pre-existing comorbidities) as potential confounders. We compared findings between two large COVID-19 patient cohorts in New York City area and Florida and demonstrated significant heterogeneity in contextual and spatial risk factors for PASC. This finding provides important implications for public health efforts to address social risk factors and help patients recover from SARS-CoV-2 infection.

Limitations of this study include: (1) we used contextual and spatial characteristics at ZCTA5 level, which may not be granular enough to estimate individuals’ exposure to risk factors. This is particularly an issue in New York City where each ZCTA5 may cover a broad geographic area and a higher number of residents. (2) Similar with many previous studies, we focused on long-term exposure to air toxicants instead of acute short-term exposure to these risk factors before SARS-CoV-2 infection.

However, previous evidence indicated that distribution of these air pollutants may have remained relatively unchanged (Chakraborty, 2021). (3) Some important potential confounders, such as vaccination status, were not adjusted due to data limitations. (4) We only included patients who sought care from the health systems affiliated with the two CRNs 31-180 days after SARS-CoV-2 infection. These patients may not be representative of patients in these two regions. (5) Patients who always tested negative might have had a positive test that was not captured in EHR (e.g., self-test at home). Thus, it is possible that some patients in the negative group may be tested positive at some point.

## 5. Conclusion

We found that multiple contextual and spatial risk factors, especially certain air pollutants and toxicants, are significantly associated with an increased risk of PASC conditions that impact multiple organ systems. These risk factors for PASC symptoms and conditions differed in the New York City area compared to Florida. Targeting interventions to reduce the burden of PASC among patients with disadvantaged contextual and spatial characteristics will help to reduce disparities of COVID-19 pandemic.

## Supporting information

Supplemental eFigures and eTables

## Data Availability

All data produced in the present work are contained in the manuscript.

## ^1^Abbreviations

PASC: post-acute sequelae of SARS-CoV-2 infection
COVID-19: the 2019 novel coronavirus disease
US: the United States
ZCTA5: 5-digit ZIP Code tabulation area
CRN: clinical research network; PCORnet, the National Patient-Centered Clinical Research Network
PM2.5: fine particulate matter with diameters that are 2.5 μm and smaller
CO: carbon monoxide
SO_2_: sulfur dioxide
NO_2_: nitrogen dioxide
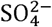: sulfate; 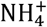 ammonium;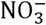^-^, nitrate
OM: organic matter
BC: black carbon
DUST: mineral dust
SS: sea-salt
O_3_: ozone
ACAG: The University of Washington at St. Louis Atmospheric Composition Analysis Group
CACES: The Center for Air, Climate, & Energy Solutions
US EPA: The United States Environmental Protection Agency
JHU CSSE: Johns Hopkins University, Center for Systems Science and Engineering Coronavirus Resource Center
CDC: The Centers for Disease Control and Prevention
NATA: National Air Toxics Assessment
USDA: US Department of Agriculture
HUD: Department of Housing and Urban Development
USPS: US Postal Service
NACIS: The North American Industry Classification System
NDVI: Normalized Difference Vegetation Index
NDI: Neighborhood Deprivation Index
ED: emergency department
VIF: variance inflation factor.

## Acknowledge

This research was funded by the National Institutes of Health (NIH) Agreement OTA HL161847-01 (contract number EHR-01-21) as part of the Researching COVID to Enhance Recovery (RECOVER) research program.

## Notes

### Competing Interest Statement

The authors have declared no competing interest.

### Author Declarations

The IRB of Weill Cornell Medicine and University of Florida gave ethical approval for this work.

## References

Al-Aly, Z., Xie, Y., & Bowe, B. (2021). High-dimensional characterization of post-acute sequelae of COVID-19. Nature, 594(7862), 259–264. https://doi.org/10.1038/s41586-021-03553-9

Bell, M. L., Catalfamo, C. J., Farland, L. V., Ernst, K. C., Jacobs, E. T., Klimentidis, Y. C., Jehn, M., & Pogreba-Brown, K. (2021). Post-acute sequelae of COVID-19 in a non-hospitalized cohort: Results from the Arizona CoVHORT. PLoS One, 16(8), e0254347. https://doi.org/10.1371/journal.pone.0254347

Bliddal, S., Banasik, K., Pedersen, O. B., Nissen, J., Cantwell, L., Schwinn, M., Tulstrup, M., Westergaard, D., Ullum, H., Brunak, S., Tommerup, N., Feenstra, B., Geller, F., Ostrowski, S. R., Gronbaek, K., Nielsen, C. H., Nielsen, S. D., & Feldt-Rasmussen, U. (2021). Acute and persistent symptoms in non-hospitalized PCR-confirmed COVID-19 patients. Sci Rep, 11(1), 13153. https://doi.org/10.1038/s41598-021-92045-x

Blomberg, B., Mohn, K. G., Brokstad, K. A., Zhou, F., Linchausen, D. W., Hansen, B. A., Lartey, S., Onyango, T. B., Kuwelker, K., Saevik, M., Bartsch, H., Tondel, C., Kittang, B. R., Bergen, C.-R. G., Cox, R. J., & Langeland, N. (2021). Long COVID in a prospective cohort of home-isolated patients. Nat Med, 27(9), 1607–1613. https://doi.org/10.1038/s41591-021-01433-3

Bull-Otterson, L., Baca, S., Saydah, S., Boehmer, T. K., Adjei, S., Gray, S., & Harris, A. M. (2022). Post– COVID Conditions Among Adult COVID-19 Survivors Aged 18–64 and≥ 65 Years—United States, March 2020–November 2021. Morbidity and Mortality Weekly Report, 71(21), 713.

Carvalho-Schneider, C., Laurent, E., Lemaignen, A., Beaufils, E., Bourbao-Tournois, C., Laribi, S., Flament, T., Ferreira-Maldent, N., Bruyere, F., Stefic, K., Gaudy-Graffin, C., Grammatico-Guillon, L., & Bernard, L. (2021). Follow-up of adults with noncritical COVID-19 two months after symptom onset. Clin Microbiol Infect, 27(2), 258–263. https://doi.org/10.1016/j.cmi.2020.09.052

Chakraborty, J. (2021). Convergence of COVID-19 and chronic air pollution risks: Racial/ethnic and socioeconomic inequities in the U.S. Environmental Research, 193. https://doi.org/ARTN11058610.1016/j.envres.2020.110586

Chen, Z., Sidell, M. A., Huang, B. Z., Chow, T., Eckel, S. P., Martinez, M. P., Gheissari, R., Lurmann, F., Thomas, D. C., Gilliland, F. D., & Xiang, A. H. (2022). Ambient Air Pollutant Exposures and COVID-19 Severity and Mortality in a Cohort of COVID-19 Patients in Southern California. Am J Respir Crit Care Med. https://doi.org/10.1164/rccm.202108-1909OC

Cohen, K., Ren, S., Heath, K., Dasmarinas, M. C., Jubilo, K. G., Guo, Y., Lipsitch, M., & Daugherty, S. E. (2022). Risk of persistent and new clinical sequelae among adults aged 65 years and older during the post-acute phase of SARS-CoV-2 infection: retrospective cohort study. BMJ, 376, e068414. https://doi.org/10.1136/bmj-2021-068414

Davis, H. E., Assaf, G. S., McCorkell, L., Wei, H., Low, R. J., Re’em, Y., Redfield, S., Austin, J. P., & Akrami, A. (2021). Characterizing long COVID in an international cohort: 7 months of symptoms and their impact. EClinicalMedicine, 38, 101019. https://doi.org/10.1016/j.eclinm.2021.101019

Diez Roux, A. V. (2001). Investigating neighborhood and area effects on health. Am J Public Health, 91(11), 1783–1789. https://doi.org/10.2105/ajph.91.11.1783

Garvin, E., Branas, C., Keddem, S., Sellman, J., & Cannuscio, C. (2013). More Than Just An Eyesore: Local Insights And Solutions on Vacant Land And Urban Health. Journal of Urban Health-Bulletin of the New York Academy of Medicine, 90(3), 412–426. https://doi.org/10.1007/s11524-012-9782-7

Groff, D., Sun, A., Ssentongo, A. E., Ba, D. M., Parsons, N., Poudel, G. R., Lekoubou, A., Oh, J. S., Ericson, J. E., Ssentongo, P., & Chinchilli, V. M. (2021). Short-term and Long-term Rates of Postacute Sequelae of SARS-CoV-2 Infection: A Systematic Review. JAMA Netw Open, 4(10), e2128568. https://doi.org/10.1001/jamanetworkopen.2021.28568

Halpin, S. J., McIvor, C., Whyatt, G., Adams, A., Harvey, O., McLean, L., Walshaw, C., Kemp, S., Corrado, J., Singh, R., Collins, T., O’Connor, R. J., & Sivan, M. (2021). Postdischarge symptoms and rehabilitation needs in survivors of COVID-19 infection: A cross-sectional evaluation. J Med Virol, 93(2), 1013–1022. https://doi.org/10.1002/jmv.26368

Hoffmann, M., Kleine-Weber, H., Schroeder, S., Kruger, N., Herrler, T., Erichsen, S., Schiergens, T. S., Herrler, G., Wu, N. H., Nitsche, A., Muller, M. A., Drosten, C., & Pohlmann, S. (2020). SARS-CoV-2 Cell Entry Depends on ACE2 and TMPRSS2 and Is Blocked by a Clinically Proven Protease Inhibitor. Cell, 181(2), 271–280 e278. https://doi.org/10.1016/j.cell.2020.02.052

Hu, H., Zheng, Y., Wen, X., Smith, S. S., Nizomov, J., Fishe, J., Hogan, W. R., Shenkman, E. A., & Bian, J. (2021). An external exposome-wide association study of COVID-19 mortality in the United States. Sci Total Environ, 768, 144832. https://doi.org/10.1016/j.scitotenv.2020.144832

Hu, M. D., Lawrence, K. G., Bodkin, M. R., Kwok, R. K., Engel, L. S., & Sandler, D. P. (2021). Neighborhood Deprivation, Obesity, and Diabetes in Residents of the US Gulf Coast. Am J Epidemiol, 190(2), 295–304. https://doi.org/10.1093/aje/kwaa206

Kaushal, R., Hripcsak, G., Ascheim, D. D., Bloom, T., Campion, T. R., Jr., Caplan, A. L., Currie, B. P., Check, T., Deland, E. L., Gourevitch, M. N., Hart, R., Horowitz, C. R., Kastenbaum, I., Levin, A. A., Low, A. F., Meissner, P., Mirhaji, P., Pincus, H. A., Scaglione, C., … Nyc, C. (2014). Changing the research landscape: the New York City Clinical Data Research Network. J Am Med Inform Assoc, 21(4), 587–590. https://doi.org/10.1136/amiajnl-2014-002764

Kim, H., Kim, W. H., Kim, Y. Y., & Park, H. Y. (2020). Air Pollution and Central Nervous System Disease: A Review of the Impact of Fine Particulate Matter on Neurological Disorders. Front Public Health, 8, 575330. https://doi.org/10.3389/fpubh.2020.575330

Kim, S. Y., Bechle, M., Hankey, S., Sheppard, L., Szpiro, A. A., & Marshall, J. D. (2020). Concentrations of criteria pollutants in the contiguous U.S., 1979 -2015: Role of prediction model parsimony in integrated empirical geographic regression. PLoS One, 15(2), e0228535. https://doi.org/10.1371/journal.pone.0228535

Kirby, J. B., Bernard, D., & Liang, L. (2021). The Prevalence of Food Insecurity Is Highest Among Americans for Whom Diet Is Most Critical to Health. Diabetes Care, 44(6), e131–e132. https://doi.org/10.2337/dc20-3116

Kurani, S. S., Lampman, M. A., Funni, S. A., Giblon, R. E., Inselman, J. W., Shah, N. D., Allen, S., Rushlow, D., & McCoy, R. G. (2021). Association Between Area-Level Socioeconomic Deprivation and Diabetes Care Quality in US Primary Care Practices. JAMA Netw Open, 4(12), e2138438. https://doi.org/10.1001/jamanetworkopen.2021.38438

Lin, W., Jiang, R., Wu, J., Wei, S., Yin, L., Xiao, X., Hu, S., Shen, Y., & Ouyang, G. (2019). Sorption properties of hydrophobic organic chemicals to micro-sized polystyrene particles. Sci Total Environ, 690, 565–572. https://doi.org/10.1016/j.scitotenv.2019.06.537

Logue, J. M., Small, M. J., & Robinson, A. L. (2011). Evaluating the national air toxics assessment (NATA): Comparison of predicted and measured air toxics concentrations, risks, and sources in Pittsburgh, Pennsylvania. Atmospheric Environment, 45(2), 476–484. https://doi.org/10.1016/j.atmosenv.2010.09.053

Mehandru, S., & Merad, M. (2022). Pathological sequelae of long-haul COVID. Nat Immunol, 23(2), 194–202. https://doi.org/10.1038/s41590-021-01104-y

Nalbandian, A., Sehgal, K., Gupta, A., Madhavan, M. V., McGroder, C., Stevens, J. S., Cook, J. R., Nordvig, A. S., Shalev, D., Sehrawat, T. S., Ahluwalia, N., Bikdeli, B., Dietz, D., Der-Nigoghossian, C., Liyanage-Don, N., Rosner, G. F., Bernstein, E. J., Mohan, S., Beckley, A. A., … Wan, E. Y. (2021). Post-acute COVID-19 syndrome. Nat Med, 27(4), 601–615. https://doi.org/10.1038/s41591-021-01283-z

Paul, K. C., Haan, M., Yu, Y., Inoue, K., Mayeda, E. R., Dang, K., Wu, J., Jerrett, M., & Ritz, B. (2020). Traffic-Related Air Pollution and Incident Dementia: Direct and Indirect Pathways Through Metabolic Dysfunction. J Alzheimers Dis, 76(4), 1477–1491. https://doi.org/10.3233/JAD-200320

Petersen, M. S., Kristiansen, M. F., Hanusson, K. D., Danielsen, M. E., B, A. S., Gaini, S., Strom, M., & Weihe, P. (2021). Long COVID in the Faroe Islands: A Longitudinal Study Among Nonhospitalized Patients. Clin Infect Dis, 73(11), e4058–e4063. https://doi.org/10.1093/cid/ciaa1792

Petroni, M., Hill, D., Younes, L., Barkman, L., Howard, S., Howell, I. B., Mirowsky, J., & Collins, M. B. (2020). Hazardous air pollutant exposure as a contributing factor to COVID-19 mortality in the United States. Environmental Research Letters, 15(9). https://doi.org/ARTN0940a910.1088/1748-9326/abaf86

Pica, N., & Bouvier, N. M. (2012). Environmental factors affecting the transmission of respiratory viruses. Curr Opin Virol, 2(1), 90–95. https://doi.org/10.1016/j.coviro.2011.12.003

Razzaq, A., Sharif, A., Aziz, N., Irfan, M., & Jermsittiparsert, K. (2020). Asymmetric link between environmental pollution and COVID-19 in the top ten affected states of US: A novel estimations from quantile-on-quantile approach. Environmental Research, 191. https://doi.org/ARTN11018910.1016/j.envres.2020.110189

Rhew, I. C., Vander Stoep, A., Kearney, A., Smith, N. L., & Dunbar, M. D. (2011). Validation of the normalized difference vegetation index as a measure of neighborhood greenness. Ann Epidemiol, 21(12), 946–952. https://doi.org/10.1016/j.annepidem.2011.09.001

Rupasingha, A., Goetz, S. J., & Freshwater, D. (2006). The production of social capital in US counties. The journal of socio-economics, 35(1), 83–101.

Sagawa, T., Tsujikawa, T., Honda, A., Miyasaka, N., Tanaka, M., Kida, T., Hasegawa, K., Okuda, T., Kawahito, Y., & Takano, H. (2021). Exposure to particulate matter upregulates ACE2 and TMPRSS2 expression in the murine lung. Environ Res, 195, 110722. https://doi.org/10.1016/j.envres.2021.110722

Shenkman, E., Hurt, M., Hogan, W., Carrasquillo, O., Smith, S., Brickman, A., & Nelson, D. (2018). OneFlorida Clinical Research Consortium: Linking a Clinical and Translational Science Institute With a Community-Based Distributive Medical Education Model. Acad Med, 93(3), 451–455. https://doi.org/10.1097/ACM.0000000000002029

Shoucri, S. M., Purpura, L., DeLaurentis, C., Adan, M. A., Theodore, D. A., Irace, A. L., Robbins-Juarez, S. Y., Khedagi, A. M., Letchford, D., Harb, A. A., Zerihun, L. M., Lee, K. E., Gambina, K., Lauring, M. C., Chen, N., Sperring, C. P., Mehta, S. S., Myers, E. L., Shih, H., … Zucker, J. E. (2021). Characterising the long-term clinical outcomes of 1190 hospitalised patients with COVID-19 in New York City: a retrospective case series. BMJ Open, 11(6), e049488. https://doi.org/10.1136/bmjopen-2021-049488

Smith, K. R., Corvalan, C. F., & Kjellstrom, T. (1999). How much global ill health is attributable to environmental factors? Epidemiology, 10(5), 573–584. https://www.ncbi.nlm.nih.gov/pubmed/10468437

Su, Y., Yuan, D., Chen, D. G., Ng, R. H., Wang, K., Choi, J., Li, S., Hong, S., Zhang, R., Xie, J., Kornilov, S. A., Scherler, K., Pavlovitch-Bedzyk, A. J., Dong, S., Lausted, C., Lee, I., Fallen, S., Dai, C. L., Baloni, P., … Heath, J. R. (2022). Multiple early factors anticipate post-acute COVID-19 sequelae. Cell, 185(5), 881–895 e820. https://doi.org/10.1016/j.cell.2022.01.014

Sudre, C. H., Murray, B., Varsavsky, T., Graham, M. S., Penfold, R. S., Bowyer, R. C., Pujol, J. C., Klaser, K., Antonelli, M., Canas, L. S., Molteni, E., Modat, M., Jorge Cardoso, M., May, A., Ganesh, S., Davies, R., Nguyen, L. H., Drew, D. A., Astley, C. M., … Steves, C. J. (2021). Attributes and predictors of long COVID. Nat Med, 27(4), 626–631. https://doi.org/10.1038/s41591-021-01292-y

Taquet, M., Dercon, Q., Luciano, S., Geddes, J. R., Husain, M., & Harrison, P. J. (2021). Incidence, co-occurrence, and evolution of long-COVID features: A 6-month retrospective cohort study of 273,618 survivors of COVID-19. PLoS Med, 18(9), e1003773. https://doi.org/10.1371/journal.pmed.1003773

Thompson, E. J., Williams, D. M., Walker, A. J., Mitchell, R. E., Niedzwiedz, C. L., Yang, T. C., Huggins, C. F., Kwong, A. S. F., Silverwood, R. J., Di Gessa, G., Bowyer, R. C. E., Northstone, K., Hou, B., Green, M. J., Dodgeon, B., Doores, K. J., Duncan, E. L., Williams, F. M. K., Open, S. C., … Steves, C. J. (2022). Long COVID burden and risk factors in 10 UK longitudinal studies and electronic health records. Nat Commun, 13(1), 3528. https://doi.org/10.1038/s41467-022-30836-0

Thomson, E. M. (2019). Air Pollution, Stress, and Allostatic Load: Linking Systemic and Central Nervous System Impacts. J Alzheimers Dis, 69(3), 597–614. https://doi.org/10.3233/JAD-190015

Tiotiu, A. I., Novakova, P., Nedeva, D., Chong-Neto, H. J., Novakova, S., Steiropoulos, P., & Kowal, K. (2020). Impact of Air Pollution on Asthma Outcomes. Int J Environ Res Public Health, 17(17). https://doi.org/10.3390/ijerph17176212

Tischer, C., Gascon, M., Fernandez-Somoano, A., Tardon, A., Lertxundi Materola, A., Ibarluzea, J., Ferrero, A., Estarlich, M., Cirach, M., Vrijheid, M., Fuertes, E., Dalmau-Bueno, A., Nieuwenhuijsen, M. J., Anto, J. M., Sunyer, J., & Dadvand, P. (2017). Urban green and grey space in relation to respiratory health in children. Eur Respir J, 49(6). https://doi.org/10.1183/13993003.02112-2015

To, T., Feldman, L., Simatovic, J., Gershon, A. S., Dell, S., Su, J., Foty, R., & Licskai, C. (2015). Health risk of air pollution on people living with major chronic diseases: a Canadian population-based study. BMJ Open, 5(9), e009075. https://doi.org/10.1136/bmjopen-2015-009075

United States Department of Agriculture. (2019). Food Environment Atlas. Retrieved 05/01 from https://www.ers.usda.gov/foodatlas/

van der Valk, J. P. M., & In ‘t Veen, J. (2021). The Interplay Between Air Pollution and Coronavirus Disease (COVID-19). J Occup Environ Med, 63(3), e163–e167. https://doi.org/10.1097/JOM.0000000000002143

van Donkelaar, A., Martin, R. V., Li, C., & Burnett, R. T. (2019). Regional Estimates of Chemical Composition of Fine Particulate Matter Using a Combined Geoscience-Statistical Method with Information from Satellites, Models, and Monitors. Environ Sci Technol, 53(5), 2595–2611. https://doi.org/10.1021/acs.est.8b06392

Walker, A. F., Hu, H., Cuttriss, N., Anez-Zabala, C., Yabut, K., Haller, M. J., & Maahs, D. M. (2020). The Neighborhood Deprivation Index and Provider Geocoding Identify Critical Catchment Areas for Diabetes Outreach. J Clin Endocrinol Metab, 105(9). https://doi.org/10.1210/clinem/dgaa462

Wang, L., Foer, D., MacPhaul, E., Lo, Y. C., Bates, D. W., & Zhou, L. (2022). PASCLex: A comprehensive post-acute sequelae of COVID-19 (PASC) symptom lexicon derived from electronic health record clinical notes. J Biomed Inform, 125, 103951. https://doi.org/10.1016/j.jbi.2021.103951

Watson, K. B., Whitfield, G. P., Thomas, J. V., Berrigan, D., Fulton, J. E., & Carlson, S. A. (2020). Associations between the National Walkability Index and walking among US Adults - National Health Interview Survey, 2015. Preventive Medicine, 137. https://doi.org/ARTN10612210.1016/j.ypmed.2020.106122

Weaver, A. K., Head, J. R., Gould, C. F., Carlton, E. J., & Remais, J. V. (2022). Environmental Factors Influencing COVID-19 Incidence and Severity. Annu Rev Public Health, 43, 271–291. https://doi.org/10.1146/annurev-publhealth-052120-101420

Wu, X., Nethery, R. C., Sabath, M. B., Braun, D., & Dominici, F. (2020). Air pollution and COVID-19 mortality in the United States: Strengths and limitations of an ecological regression analysis. Sci Adv, 6(45). https://doi.org/10.1126/sciadv.abd4049

Xie, Y., Bowe, B., & Al-Aly, Z. (2021). Burdens of post-acute sequelae of COVID-19 by severity of acute infection, demographics and health status. Nat Commun, 12(1), 6571. https://doi.org/10.1038/s41467-021-26513-3

Yoo, S. M., Liu, T. C., Motwani, Y., Sim, M. S., Viswanathan, N., Samras, N., Hsu, F., & Wenger, N. S. (2022). Factors Associated with Post-Acute Sequelae of SARS-CoV-2 (PASC) After Diagnosis of Symptomatic COVID-19 in the Inpatient and Outpatient Setting in a Diverse Cohort. J Gen Intern Med. https://doi.org/10.1007/s11606-022-07523-3

Zang, C., Zhang, Y., Xu, J., Bian, J., Morozyuk, D., Schenck, E., Khullar, D., Nordvig, A. S., Shenkman, E., Rothman, R. L., Block, J. P., Lyman, K., Weiner, M., Carton, T. W., Wang, F., & Kaushal, R. (2022). Understanding Post-Acute Sequelae of SARS-CoV-2 Infection through Data-Driven Analysis with Longitudinal Electronic Health Records: Findings from the RECOVER Initiative. https://doi.org/medRxiv doi: https://doi.org/10.1101/2022.05.21.22275420

Zhang, Y., Khullar, D., Wang, F., Steel, P., Wu, Y., Orlander, D., Weiner, M., & Kaushal, R. (2021). Socioeconomic variation in characteristics, outcomes, and healthcare utilization of COVID-19 patients in New York City. PLoS One, 16(7), e0255171. https://doi.org/10.1371/journal.pone.0255171

Zhang, Z., Weichenthal, S., Kwong, J. C., Burnett, R. T., Hatzopoulou, M., Jerrett, M., van Donkelaar, A., Bai, L., Martin, R. V., Copes, R., Lu, H., Lakey, P., Shiraiwa, M., & Chen, H. (2021). A Population-Based Cohort Study of Respiratory Disease and Long-Term Exposure to Iron and Copper in Fine Particulate Air Pollution and Their Combined Impact on Reactive Oxygen Species Generation in Human Lungs. Environ Sci Technol, 55(6), 3807–3818. https://doi.org/10.1021/acs.est.0c05931

Zhou, X., Josey, K., Kamareddine, L., Caine, M. C., Liu, T., Mickley, L. J., Cooper, M., & Dominici, F. (2021). Excess of COVID-19 cases and deaths due to fine particulate matter exposure during the 2020 wildfires in the United States. Sci Adv, 7(33). https://doi.org/10.1126/sciadv.abi8789

