## Supplemental eFigures and eTables for "Identifying Contextual and Spatial Risk Factors for Post-Acute Sequelae of SARS-CoV-2 Infection: An EHR-based Cohort Study from the RECOVER Program"

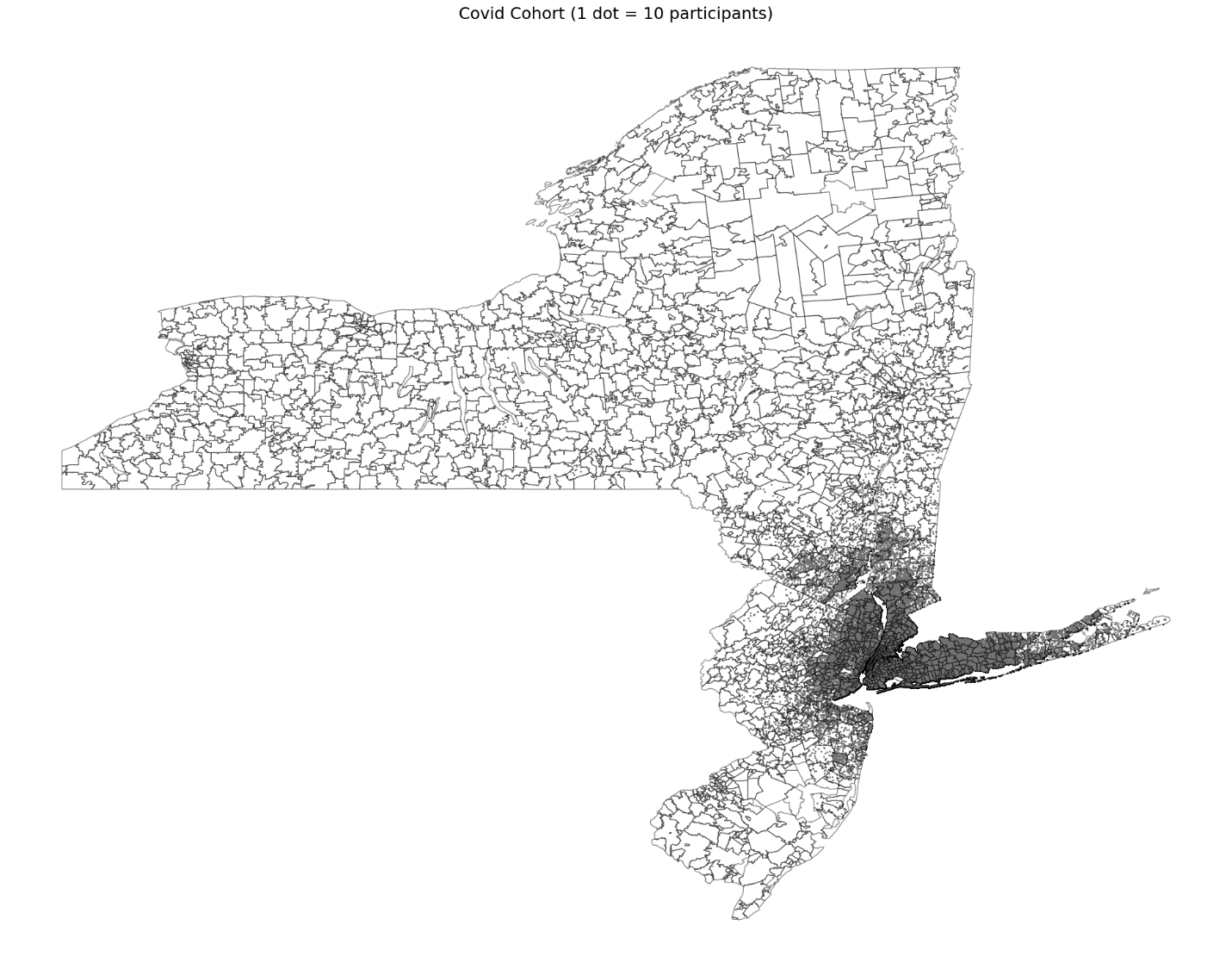

### **eFigure 1 Catchment Area of COVID-19 Patients in INSIGHT CRN, All ZCTA5**

### **eFigure 2 Catchment Area of COVID-19 Patients in OneFlorida+ CRN**

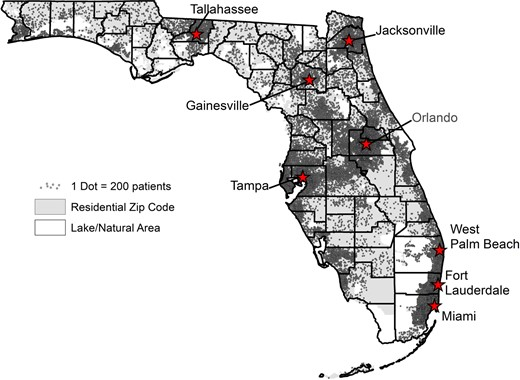

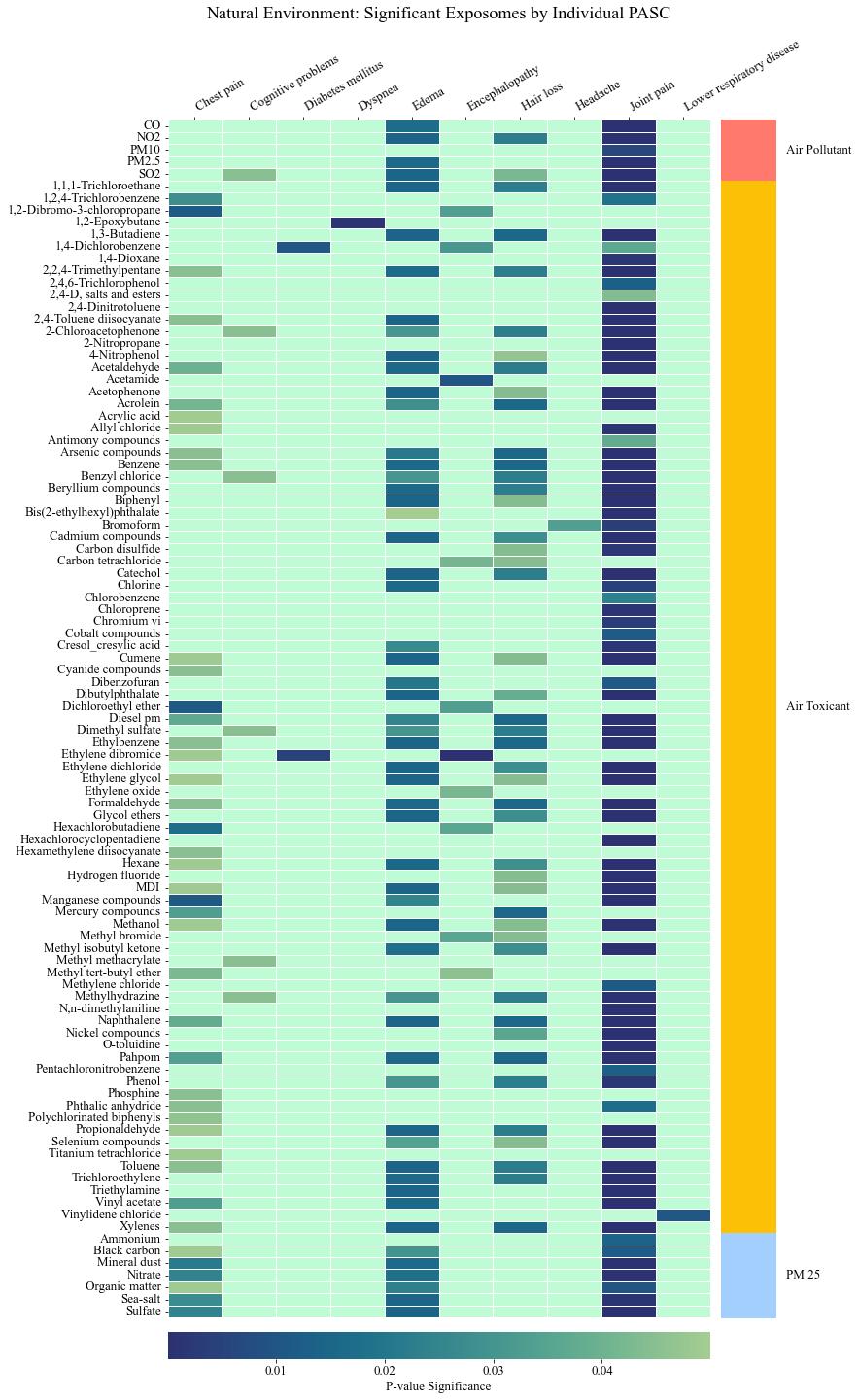

### **eFigure 3a Significant Contextual and Spatial Factors Associated with Individual PASC Conditions in Phase 1 Analysis Using INSIGHT Sample: Nature Environment Factors**

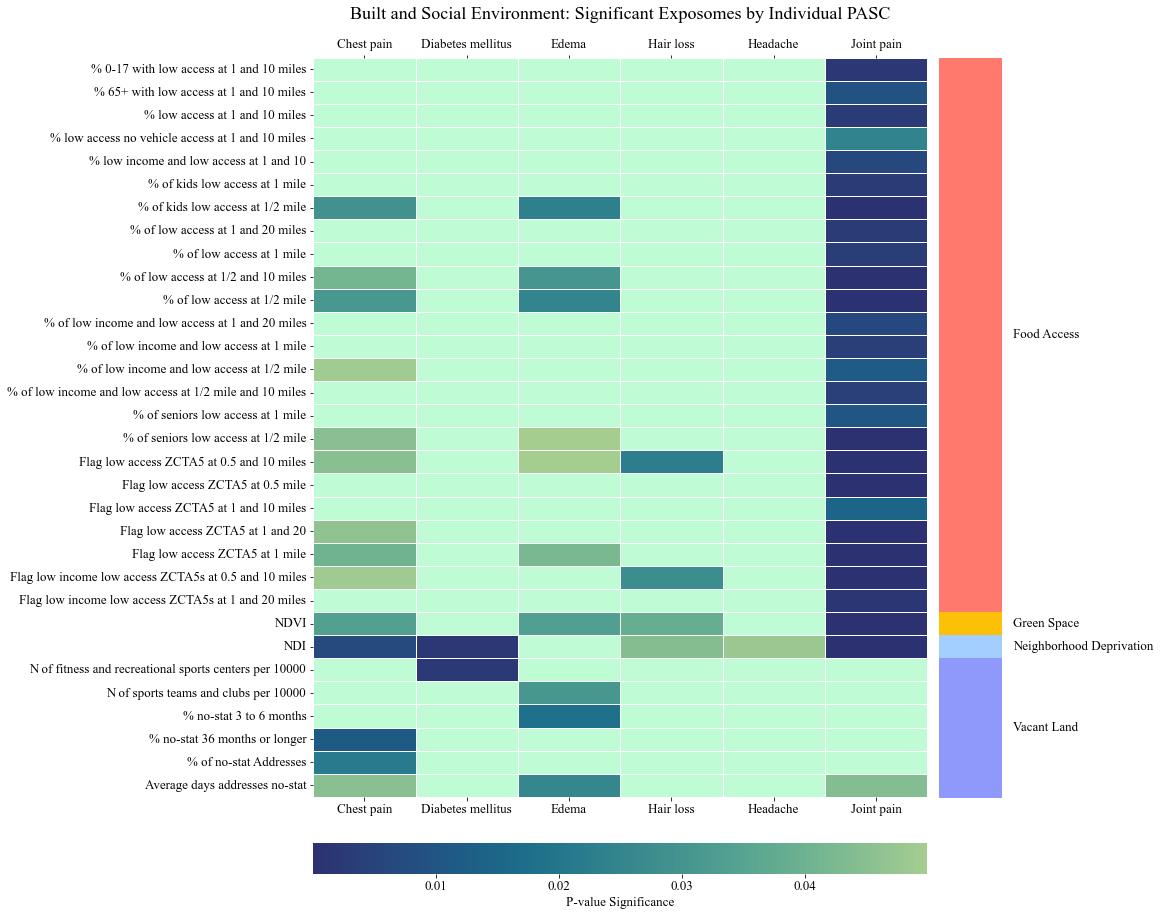

### **eFigure 3b Significant Contextual and Spatial Factors Associated with Individual PASC Conditions in Phase 1 Analysis Using INSIGHT Sample: Built Environment and Social Environment Factors**

Notes: Figure represent contextual and spatial characteristics identified from mixed effects logistic regressions where a PASC condition is the outcome and each contextual and spatial characteristics characteristic is the key independent variable. All regressions controlled for patient-level covariates. A contextual and spatial characteristic is considered significant if the false discovery rate adjusted p value is < 0.05.

### **eFigure 4a Significant Contextual and Spatial Factors Associated with Individual PASC Conditions in Phase 1 Analysis Using OneFlorida+ Sample: Nature Environment Factors**

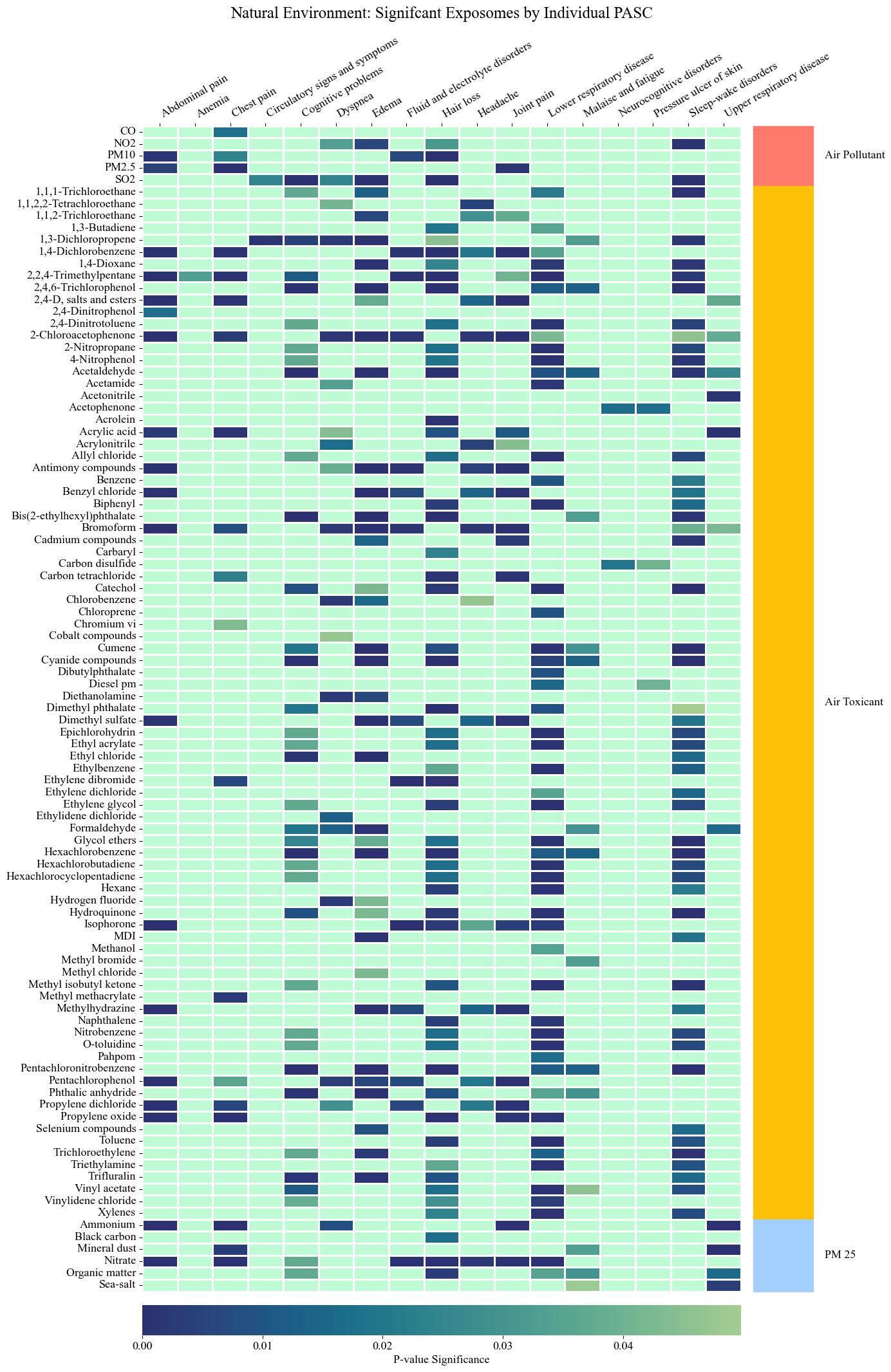

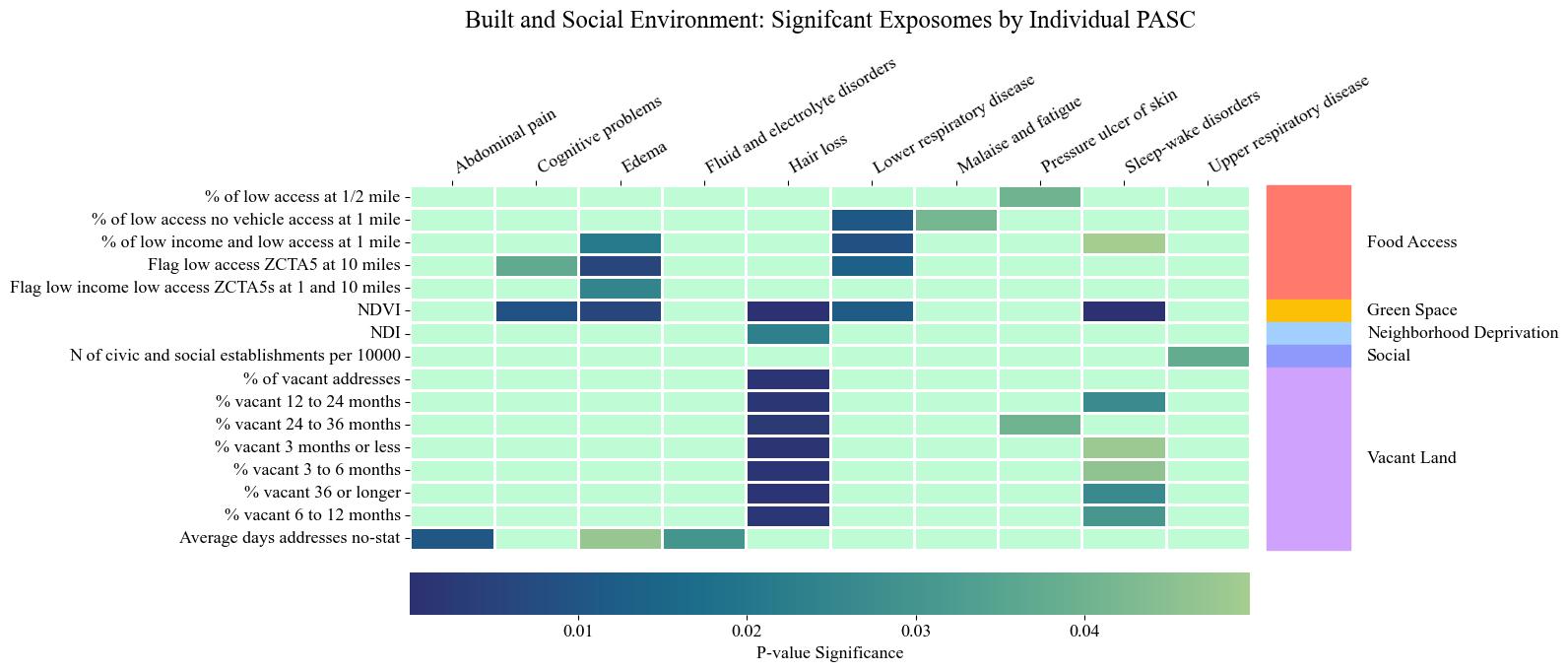

### **eFigure 4b Significant Contextual and Spatial Factors Associated with Individual PASC Conditions in Phase 1 Analysis Using OneFlorida+ Sample: Built Environment and Social Environment Factors**

Notes: Figure represent contextual and spatial characteristics identified from mixed effects logistic regressions where a PASC condition is the outcome and each contextual and spatial characteristics characteristic is the key independent variable. All regressions controlled for patient-level covariates. A contextual and spatial characteristic is considered significant if the false discovery rate adjusted p value is < 0.05.

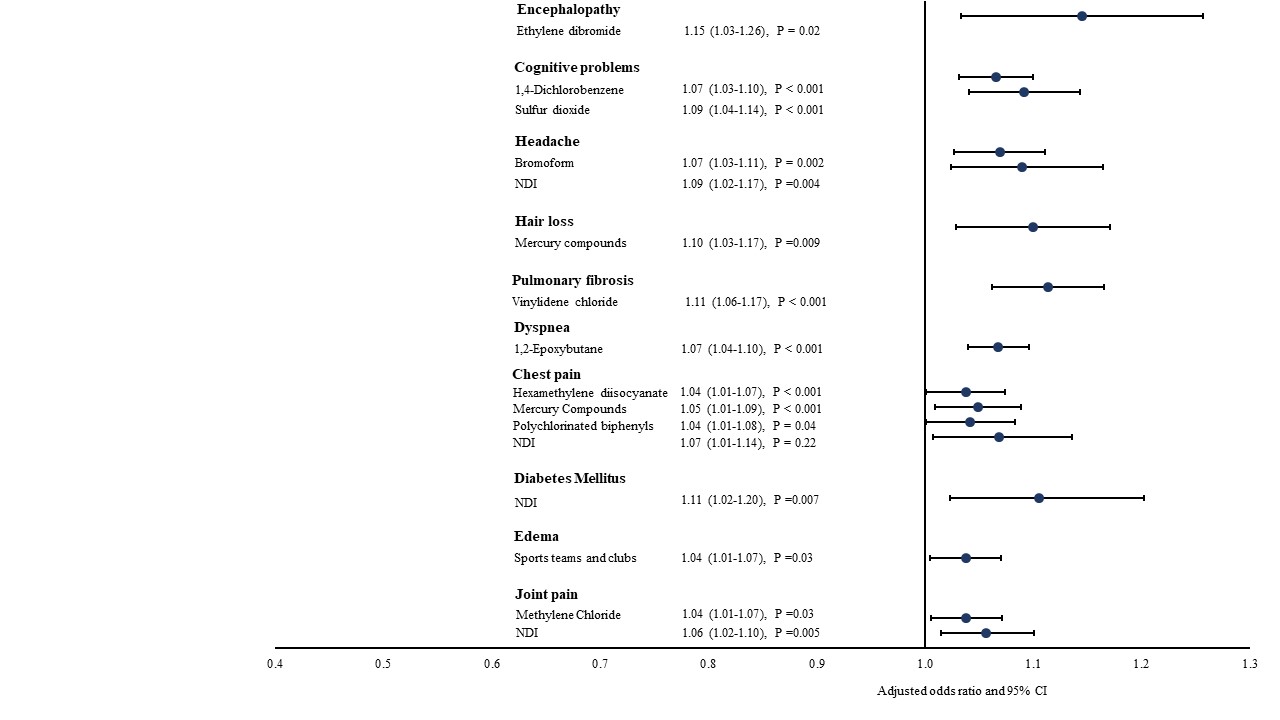

### **eFigure 5 Contextual and Spatial Risk Factors for Individual PASC Conditions using INSIGHT Sample**

Notes: NDI: Neighborhood Deprivation Index**.** ORs were estimated from mixed effects logistic regressions with ZCTA5 random intercept. Each regression includes all significant neighborhood and environmental characteristics identified from phase 1 analysis for each PASC outcome, controlling for all patient-level covariates.

### **eFigure 6 Contextual and Spatial Risk Factors for Individual PASC Conditions using OneFlorida+ Sample**

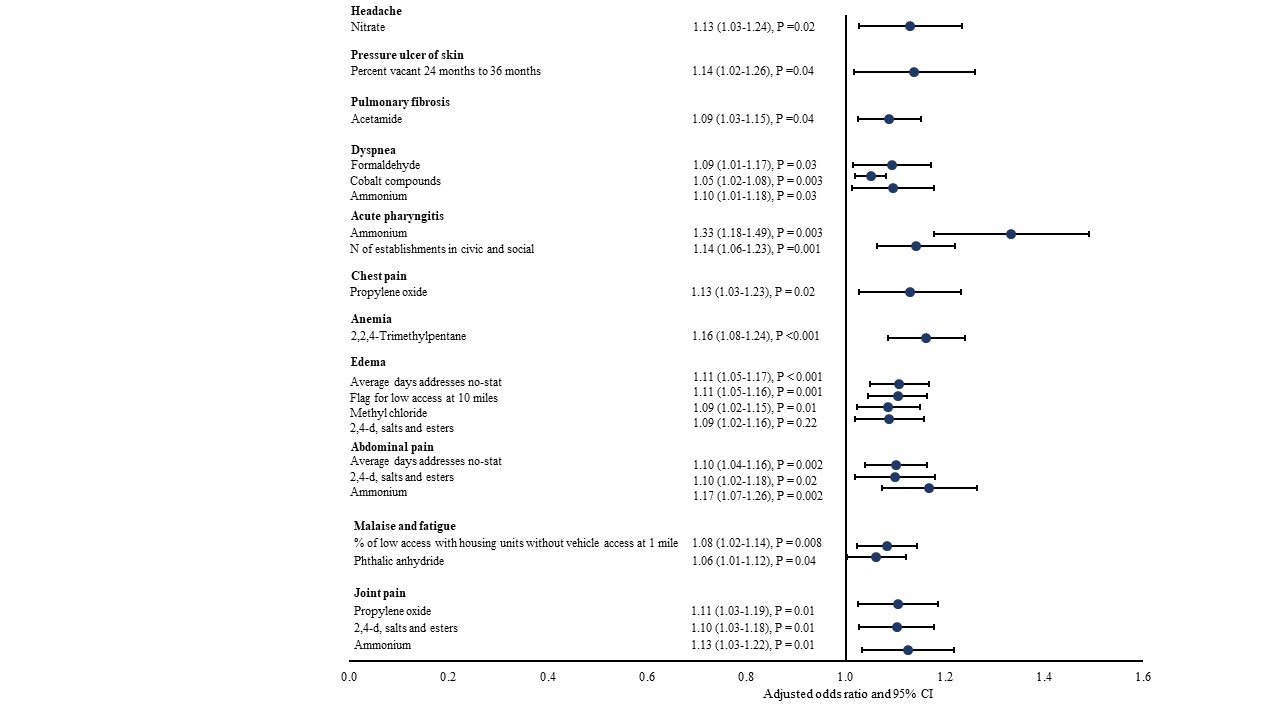

Notes: NDI: Neighborhood Deprivation Index**.** ORs were estimated from mixed effects logistic regressions with ZCTA5 random intercept. Each regression includes all significant neighborhood and environmental characteristics identified from phase 1 analysis for each PASC outcome, controlling for all patient-level covariates.

### **eTable 1 All Neighborhood and Environmental Characteristics Considered in the Study**

| Variables | Source | Category |
| --- | --- | --- |
| Average Days Addresses Vacant | USHUD | Vacant land |
| Average days addresses no-stat | USHUD |  |
| Percent of vacant addresses | USHUD |  |
| Percent vacant 3 months to less | USHUD |  |
| Percent vacant 3 months to 6 months | USHUD |  |
| Percent vacant 6 months to 12 months | USHUD |  |
| Percent vacant 12 months to 24 months | USHUD |  |
| Percent vacant 24 months to 36 months | USHUD |  |
| Percent vacant 36 months or longer | USHUD |  |
| Percent vacant 36 months or longer | USHUD |  |
| Percent of no-stat Addresses | USHUD |  |
| Percent no-stat 3 months to less | USHUD |  |
| Percent no-stat 3 months to 6 months | USHUD |  |
| Percent no-stat 6 months to 12 months | USHUD |  |
| Percent no-stat 12 months to 24 months | USHUD |  |
| Percent no-stat 24 months to 36 months | USHUD |  |
| Percent no-stat 36 months or longer | USHUD |  |
| Percent previous quarter no-stat currently in service | USHUD |  |
| 1,1,1-trichloroethane | NATA | Air toxicants |
| 1,1,2-trichloroethane | NATA |  |
| 1,1,2,2-tetrachloroethane | NATA |  |
| 1,2-dibromo-3-chloropropane | NATA |  |
| 1,2-epoxybutane | NATA |  |
| 1,2,4-trichlorobenzene | NATA |  |
| 1,3-butadiene | NATA |  |
| 1,3-dichloropropene | NATA |  |
| 1,3-propane sultone | NATA |  |
| 1,4-dichlorobenzene | NATA |  |
| 1,4-dioxane | NATA |  |
| 2-chloroacetophenone | NATA |  |
| 2-nitropropane | NATA |  |
| 2,2,4-trimethylpentane | NATA |  |
| 2,4-d, salts and esters | NATA |  |
| 2,4-dinitrophenol | NATA |  |
| 2,4-dinitrotoluene | NATA |  |
| 2,4-toluene diisocyanate | NATA |  |
| 2,4,6-trichlorophenol | NATA |  |
| 4-nitrophenol | NATA |  |
| 4,4'-methylene bis(2-chloroaniline) | NATA |  |
| 4,4'-methylenedianiline | NATA |  |
| 4,4'-methylenediphenyl diisocyanate (mdi) | NATA |  |
| 4,6-dinitro-o-cresol (including salts) | NATA |  |
| Acetaldehyde | NATA |  |
| Acetamide | NATA |  |
| Acetonitrile | NATA |  |
| Acetophenone | NATA |  |
| Acrolein | NATA |  |
| Acrylamide | NATA |  |
| Acrylic acid | NATA |  |
| Acrylonitrile | NATA |  |
| Allyl chloride | NATA |  |
| Aniline | NATA |  |
| Anisidine | NATA |  |
| Antimony compounds | NATA |  |
| Arsenic compounds (inorganic including arsine) | NATA |  |
| Benzene | NATA |  |
| Benzyl chloride | NATA |  |
| Beryllium compounds | NATA |  |
| Biphenyl | NATA |  |
| Bis(2-ethylhexyl) phthalate (dehp) | NATA |  |
| Bromoform | NATA |  |
| Cadmium compounds | NATA |  |
| Captan | NATA |  |
| Carbaryl | NATA |  |
| Carbon disulfide | NATA |  |
| Carbon tetrachloride | NATA |  |
| Carbonyl sulfide | NATA |  |
| Catechol | NATA |  |
| Chlordane | NATA |  |
| Chlorine | NATA |  |
| Chloroacetic acid | NATA |  |
| Chlorobenzene | NATA |  |
| Chloroform | NATA |  |
| Chloroprene | NATA |  |
| Chromium vi (hexavalent) | NATA |  |
| Cobalt compounds | NATA |  |
| Coke oven emissions | NATA |  |
| Cresol cresylic acid (mixed isomers) | NATA |  |
| Cumene | NATA |  |
| Cyanide compounds | NATA |  |
| Dibenzofuran | NATA |  |
| Dibutyl phthalate | NATA |  |
| Dichloroethyl ether | NATA |  |
| Dichlorvos | NATA |  |
| Diesel pm | NATA |  |
| Diethanolamine | NATA |  |
| Dimethyl formamide | NATA |  |
| Dimethyl phthalate | NATA |  |
| Dimethyl sulfate | NATA |  |
| Epichlorohydrin | NATA |  |
| Ethyl acrylate | NATA |  |
| Ethyl carbamate (urethane) chloride (chloroethane) | NATA |  |
| Ethyl chloride | NATA |  |
| Ethylbenzene | NATA |  |
| Ethylene dibromide (dibromoethane) | NATA |  |
| Ethylene dichloride (1,2-dichloroethane) | NATA |  |
| Ethylene glycol | NATA |  |
| Ethylene oxide | NATA |  |
| Ethylene thiourea | NATA |  |
| Ethylidene dichloride (1,1-dichloroethane) | NATA |  |
| Formaldehyde | NATA |  |
| Glycol ethers | NATA |  |
| Heptachlor | NATA |  |
| Hexachlorobenzene | NATA |  |
| Hexachlorobutadiene | NATA |  |
| Hexachlorocyclopentadiene | NATA |  |
| Hexamethylene diisocyanate | NATA |  |
| Hexane | NATA |  |
| Hydrazine | NATA |  |
| Hydrochloric acid (hydrogen chloride [gas only]) | NATA |  |
| Hydrogen fluoride (hydrofluoric acid) | NATA |  |
| Hydroquinone | NATA |  |
| Isophorone | NATA |  |
| Lead compounds | NATA |  |
| Maleic anhydride | NATA |  |
| Manganese compounds | NATA |  |
| Mercury compounds | NATA |  |
| Methanol | NATA |  |
| Methyl bromide (bromomethane) | NATA |  |
| Methyl chloride (chloromethane) | NATA |  |
| Methyl iodide (iodomethane) | NATA |  |
| Methyl isobutyl ketone (hexone) | NATA |  |
| Methyl methacrylate | NATA |  |
| Methyl tert-butyl ether | NATA |  |
| Methylene chloride | NATA |  |
| Methylhydrazine | NATA |  |
| N,n-dimethylaniline | NATA |  |
| Naphthalene | NATA |  |
| Nickel compounds | NATA |  |
| Nitrobenzene | NATA |  |
| O-toluidine | NATA |  |
| P-phenylenediamine | NATA |  |
| Pahpom | NATA |  |
| Pentachloronitrobenzene (quintobenzene) | NATA |  |
| Pentachlorophenol | NATA |  |
| Phenol | NATA |  |
| Phosgene | NATA |  |
| Phosphine | NATA |  |
| Phosphorus | NATA |  |
| Phthalic anhydride | NATA |  |
| Polychlorinated biphenyls (aroclors) | NATA |  |
| Propionaldehyde | NATA |  |
| Propylene dichloride (1,2-dichloropropane) | NATA |  |
| Propylene oxide | NATA |  |
| Quinoline | NATA |  |
| Quinone (p-benzoquinone) | NATA |  |
| Selenium compounds | NATA |  |
| Styrene | NATA |  |
| Tetrachloroethylene | NATA |  |
| Titanium tetrachloride | NATA |  |
| Toluene | NATA |  |
| Trichloroethylene | NATA |  |
| Triethylamine | NATA |  |
| Trifluralin | NATA |  |
| Vinyl acetate | NATA |  |
| Vinyl chloride | NATA |  |
| Vinylidene chloride | NATA |  |
| Xylenes (mixed isomers) | NATA |  |
| Percentage of low access population at 1 mile for urban and 10 miles for rural | FARA | Food Access |
| Percentage of low income and low access population at 1 mile for urban and 10 miles for rural | FARA |  |
| Percentage of low access population with housing units without vehicle access at 1 mile for urban and 10 miles for rural | FARA |  |
| Percentage of children age 0-17 with low access at 1 mile for urban and 10 miles for rural | FARA |  |
| Percentage of seniors age 65+ with low access at 1 mile for urban and 10 miles for rural | FARA |  |
| Flag for low-income low access ZCTA5s at 1 mile for urban and 10 miles for rural | FARA |  |
| Flag for low access ZCTA5 at 1 mile for urban areas and 10 miles for rural areas | FARA |  |
| Flag for low-income low access ZCTA5s at 0.5 mile for urban and 10 miles for rural | FARA |  |
| Flag for low-income low access ZCTA5s at 1 mile for urban and 20 miles for rural | FARA |  |
| Flag for low access ZCTA5 at 0.5 mile for urban and 10 miles for rural | FARA |  |
| Flag for low access ZCTA5 at 1 mile for urban and 20 miles for rural | FARA |  |
| Flag for low access ZCTA5 at 0.5 mile | FARA |  |
| Flag for low access ZCTA5 at 1 mile | FARA |  |
| Flag for low access ZCTA5 at 10 miles | FARA |  |
| Flag for low access ZCTA5 at 20 miles | FARA |  |
| Flag for low access ZCTA5 using vehicle access and at 20 miles in rural areas | FARA |  |
| Flag for ZCTA5 where >= 100 of households do not have a vehicle, and beyond 1/2 mile from supermarket | FARA |  |
| Group quarters, ZCTA5 with high share (>=67%) | FARA |  |
| Percent of ZCTA5 population residing in group quarters | FARA |  |
| Percentage of low access population at 1/2 mile | FARA |  |
| Percentage of low access population that are low income at 1/2 mile | FARA |  |
| Percentage of low access population that are kids at 1/2 mile | FARA |  |
| Percentage of low access population that are seniors at 1/2 mile | FARA |  |
| Percentage of low access population with housing units without vehicle access at 1/2 mile | FARA |  |
| Percentage of low access population at 1 mile | FARA |  |
| Percentage of low access population that are low income at 1 mile | FARA |  |
| Percentage of low access population that are kids at 1 mile | FARA |  |
| Percentage of low access population that are seniors at 1 mile | FARA |  |
| Percentage of low access population with housing units without vehicle access at 1 mile | FARA |  |
| Percentage of low access population at 10 miles | FARA |  |
| Percentage of low access population that are low income at 10 miles | FARA |  |
| Percentage of low access population that are kids at 10 miles | FARA |  |
| Percentage of low access population that are seniors at 10 miles | FARA |  |
| Percentage of low access population with housing units without vehicle access at 10 miles | FARA |  |
| Percentage of low access population at 20 miles | FARA |  |
| Percentage of low access population that are low income at 20 miles | FARA |  |
| Percentage of low access population that are kids at 20 miles | FARA |  |
| Percentage of low access population that are seniors at 20 miles | FARA |  |
| Percentage of low access population with housing units without vehicle access at 20 miles | FARA |  |
| Percentage of low access population at 1/2 mile for urban and 10 miles for rural | FARA |  |
| Percentage of low access population at 1 mile for urban and 20 miles for rural | FARA |  |
| Percentage of low income and low access population at 1/2 mile for urban and 10 miles for rural | FARA |  |
| Percentage of low income and low access population at 1 mile for urban and 20 miles for rural | FARA |  |
| CO (ppm) | CACES | Criteria air pollutants |
| NO2 (ppb) | CACES |  |
| O3 (ppb) | CACES |  |
| PM10 (µg/m3) | CACES |  |
| PM2.5 (µg/m3) | CACES |  |
| SO2 (ppb) | CACES |  |
| National Walkability Index | WI | Walkability |
| Number of establishments in religious organizations per 10000 population | CBP | Social capital |
| Number of establishments in civic and social associations per 10000 population | CBP |  |
| Number of establishments in business associations per 10000 population | CBP |  |
| Number of establishments in political organizations per 10000 population | CBP |  |
| Number of establishments in professional organizations per 10000 population | CBP |  |
| Number of establishments in labor organizations per 10000 population | CBP |  |
| Number of establishments in bowling center per 10000 population | CBP |  |
| Number of establishments in fitness and recreational sports centers per 10000 population | CBP |  |
| Number of establishments in gold courses and country clubs per 10000 population | CBP |  |
| Number of establishments in sports teams and clubs per 10000 population | CBP |  |
| Black carbon (BC) | ACAG | PM2.5 compositions |
| Ammonium (NH4) | ACAG |  |
| Nitrate (NO3) | ACAG |  |
| Organic matter (OM) | ACAG |  |
| Sulfate (SO4) | ACAG |  |
| Mineral dust (DUST) | ACAG |  |
| Sea-salt (SS) | ACAG |  |
| NDVI | NDVI | Green Space |
| NDI | NDI | Neighborhood Deprivation |

Notes: ACAG: Atmospheric Composition Analysis Group; CACES: Center for Air, Climate, & Energy Solutions; EPA: Environmental Protection Agency; NATA: National Air Toxics Assessment; HUD: Department of Housing and Urban Development; USDA: US Department of Agriculture; FARA: Food Access Research Atlas; NASA: National Aeronautics and Space Administration; MODIS: Moderate Resolution Imaging Spectroradiometer; ACS: American Community Survey; CBP: Census Business Pattern; UCR: Uniform Crime Reporting. NDVI: Normalized Difference Vegetation Index; NDI: Neighborhood Deprivation Index.

### **eTable 2 Excluded Neighborhood and Environmental Characteristics in Both CRNs**

| lINSIGHT | OneFlorida+ |
| --- | --- |
| Flag for ZCTA5 where >= 100 of households do not have a vehicle, and beyond 1/2 mile from supermarket | Flag for ZCTA5 where >= 100 of households do not have a vehicle, and beyond 1/2 mile from supermarket |
| Flag for low access ZCTA5 at 10 miles | Ethylene thiourea |
| Flag for low access ZCTA5 at 20 miles | Heptachlor |
| Flag for low access ZCTA5 using vehicle access and at 20 miles in rural areas | 1,3-propane sultone |
| Flag for low-income low access ZCTA5s at 1 mile for urban and 10 miles for rural | P-phenylenediamine |
| Percentage of low access population with housing units without vehicle access at 10 miles | Phosgene |
| Percentage of low access population with housing units without vehicle access at 20 miles | Phosphine |
| Percentage of low access population that are kids at 10 miles | Quinoline |
| Percentage of low access population that are kids at 20 miles | Titanium tetrachloride |
| Percentage of low access population that are low income at 10 miles | 4,4'-methylene bis(2-chloroaniline) |
| Percentage of low access population that are low income at 20 miles | 4,4'-methylenedianiline |
| Percentage of low access population at 10 miles | 4,6-dinitro-o-cresol (including salts) |
| Percentage of low access population at 20 miles | Acrylamide |
| Percentage of low access population that are seniors at 10 miles | Aniline |
| Percentage of low access population that are seniors at 20 miles | Anisidine |
| Ethylene thiourea | 1,2-dibromo-3-chloropropane |
| Heptachlor | Chlordane |
| Methyl iodide (iodomethane) | 1,2-epoxybutane |
| 1,3-propane sultone | Chloroacetic acid |
| P-phenylenediamine | Coke oven emissions |
| Phosgene | Dichloroethyl ether (bis[2-chloroethyl]ether) |
| Quinone (p-benzoquinone) | Dichlorvos |
| 4,4'-methylene bis(2-chloroaniline) | Ethyl carbamate (urethane) chloride (chloroethane) |
| 4,4'-methylenedianiline | Aggravated assault rate |
| 4,6-dinitro-o-cresol (including salts) | Arson rate |
| Acrylamide | Burglary rate |
| Anisidine | Curfew and loitering rate |
| Chlordane | Disorderly conduct rate |
| Chloroacetic acid | Drug abuse rate |
| Coke oven emissions | Sale of drug rate |
| Ethyl carbamate (urethane) chloride (chloroethane) | Drunkenness rate |
| Aggravated assault rate | Driving under the influence rate |
| Arson rate | Embezzlement rate |
| Burglary rate | Offense against the family and children rate |
| Curfew and loitering rate | Forgery rate |
| Disorderly conduct rate | Fraud rate |
| Drug abuse rate | Gambling rate |
| Sale of drug rate | Violation of liquor laws rate |
| Drunkenness rate | Manslaughter rate |
| Driving under the influence rate | Motor vehicle theft rate |
| Embezzlement rate | Murder rate |
| Offense against the family and children rate | Other assault rate |
| Forgery rate | Other sex offense rate |
| Fraud rate | Other crime rate |
| Gambling rate | Possession of drug rate |
| Violation of liquor laws rate | Prostitution rate |
| Manslaughter rate | Rape rate |
| Motor vehicle theft rate | Robbery rate |
| Murder rate | Runaways rate |
| Other assault rate | Stolen property rate |
| Other sex offense rate | Suspicion rate |
| Other crime rate | Theft rate |
| Possession of drug rate | Vagrancy rate |
| Prostitution rate | Vandalism rate |
| Rape rate | Violation of weapon carrying or possessing rate |
| Robbery rate |  |
| Runaways rate |  |
| Stolen property rate |  |
| Suspicion rate |  |
| Theft rate |  |
| Vagrancy rate |  |
| Vandalism rate |  |
| Violation of weapon carrying or possessing rate |  |

### **eTable 3 Interactions between Contextual and Spatial Risk Factors and PASC by Organ System, Analysis Using INSIGHT Sample**

| **PASC Outcomes** | **Key Independ Variables** | **Adjusted ORs** | **P-value** |
| --- | --- | --- | --- |
| Any nervous PASC | COVID-19 indicator (1=positive) | 1.32 | <0.001 |
|  | Methyl methacrylate | 1.02 | 0.09 |
|  | Interaction between methyl methacrylate and COVID-19 | 1.02 | 0.29 |
|  | N of civic and social establishments per 10000 | 1.02 | 0.13 |
|  | Interaction between N of civic and social establishments per 10000 and COVID-19 | 1.04 | 0.02 |
| Any skin PASC | COVID-19 indicator (1=positive) | 1.67 | <0.001 |
|  | Mercury compounds | 1.06 | 0.009 |
|  | Interaction between mercury compounds and COVID-19 | 1.03 | 0.47 |
| Any respiratory PASC | COVID-19 indicator (1=positive) | 1.81 | <0.001 |
|  | 1,2-Epoxybutane | 1.00 | 0.76 |
|  | Interaction between 1,2-Epoxybutane and COVID-19 | 1.07 | <0.001 |
| Any endocrine PASC | COVID-19 indicator (1=positive) | 1.16 | <0.001 |
|  | NDI | 1.12 | <0.001 |
|  | Interaction between NDI and COVID-19 | 0.96 | 0.03 |

Notes: NDI: neighborhood deprivation index.

### **eTable 4 Interactions between Contextual and Spatial Risk Factors and Individual PASC, Analysis Using INSIGHT Sample**

| **PASC Outcomes** | **Key Independ Variables** | **Adjusted ORs** | **P-value** |
| --- | --- | --- | --- |
| Encephalopathy | COVID-19 indicator (1=positive) | 1.45 | <0.001 |
|  | Ethylene dibromide | 1.02 | 0.40 |
|  | Interaction between Ethylene dibromide and COVID-19 | 1.13 | <0.001 |
| Cognitive problems | COVID-19 indicator (1=positive) | 1.31 | <0.001 |
|  | 1,4-Dichlorobenzene | 0.99 | 0.009 |
|  | Interaction between 1,4-Dichlorobenzene and COVID-19 | 1.06 | 0.007 |
|  | Sulfur dioxide | 1.15 | <0.001 |
|  | Interaction between sulfur dioxide and COVID-19 | 0.94 | 0.03 |
| Headache | COVID-19 indicator (1=positive) | 1.30 | <0.001 |
|  | Bromoform | 1.07 | <0.001 |
|  | Interaction between bromoform and COVID-19 | 0.99 | 0.73 |
|  | NDI | 1.08 | <0.001 |
|  | Interaction between NDI and COVID-19 | 0.95 | 0.09 |
| Hair loss | COVID-19 indicator (1=positive) | 1.97 | <0.001 |
|  | Mercury compounds | 1.08 | 0.01 |
|  | Interactions between mercury compounds and COVID-19 | 1.04 | 0.42 |
| Pulmonary fibrosis | COVID-19 | 2.04 | <0.001 |
|  | Vinylidene chloride | 1.07 | 0.001 |
|  | Interaction between vinylidene chloride and COVID-19 | 1.04 | 0.18 |
| Dyspnea | COVID-19 | 1.81 | <0.001 |
|  | 1,2-Epoxybutane | 1.00 | 0.77 |
|  | Interaction between 1,2-Epoxybutane and COVID-19 | 1.08 | <0.001 |
| Chest pain | COVID-19 | 1.51 | <0.001 |
|  | Hexamethylene diisocyanate | 1.03 | 0.06 |
|  | Interaction between hexamethylene diisocyanate and COVID-19 | 1.01 | 0.63 |
|  | Mercury compounds | 1.003 | 0.84 |
|  | Interaction between mercury compounds and COVID-19 | 1.07 | 0.03 |
|  | Polychlorinated biphenyls | 0.93 | <0.001 |
|  | Interaction between polychlorinated biphenyls and COVID-19 | 1.16 | <0.001 |
|  | NDI | 1.14 | <0.001 |
|  | Interaction between NDI and COVID-19 | 0.88 | <0.001 |
| Diabetes Mellitus | COVID-19 | 1.17 | <0.001 |
|  | NDI | 1.11 | <0.001 |
|  | Interaction between NDI and COVID-19 | 0.96 | 0.28 |
| Edema | COVID-19 | 1.11 | <0.001 |
|  | Sports teams and clubs | 1.01 | 0.59 |
|  | Interaction between sports teams and clubs and COVID-19 | 1.04 | 0.04 |
| Joint pain | COVID-19 | 1.15 | <0.001 |
|  | Methylene chloride | 1.02 | 0.13 |
|  | Interaction between COVID-19 and methylene Chloride | 1.02 | 0.28 |
|  | NDI | 1.13 | <0.001 |
|  | Interaction between NDI and COVID-19 | 0.98 | 0.21 |

Notes: NDI: neighborhood deprivation index

### **eTable 5 Interactions between Contextual and Spatial Risk Factors and PASC by Organ System, Analysis Using OneFlorida+ Sample**

| **PASC Outcomes** | **Key Independ Variables** | **Adjusted ORs** | **P-value** |
| --- | --- | --- | --- |
| Any nervous PASC | COVID-19 indicator (1=positive) | 0.75 | <0.001 |
|  | Nitrate | 1.00 | 0.98 |
|  | Interaction between nitrate and COVID-19 | 1.19 | <0.001 |
| Any skin PASC | COVID-19 indicator (1=positive) | 1.84 | <0.001 |
|  | Mercury compounds | 0.87 | <0.001 |
|  | Interaction between mercury compounds and COVID-19 | 0.91 | 0.27 |
| Any respiratory PASC | COVID-19 indicator (1=positive) | 0.98 | 0.65 |
|  | Formaldehyde | 1.03 | 0.43 |
|  | Interaction between formaldehyde e and COVID-19 | 1.17 | 0.004 |
|  | Ethylene dibromide | 1.02 | 0.27 |
|  | Interaction between ethylene dibromide and COVID-19 | 0.78 | <0.001 |
| Any circulatory PASC | COVID-19 indicator (1=positive) | 0.80 | <0.001 |
|  | Ammonium (NH4) | 1.01 | 0.78 |
|  | Interaction between NH4 and COVID-19 | 1.28 | <0.001 |
| Any blood PASC | COVID-19 indicator (1=positive) | 0.82 | <0.001 |
|  | 2,2,4-Trimethylpentane | 1.19 | <0.001 |
|  | Interaction between 2,2,4-Trimethylpentane and COVID-19 | 1.01 | 0.83 |
| Any endocrine PASC | COVID-19 indicator (1=positive) | 0.68 | <0.001 |
|  | Average days addresses no-stat | 1.07 | <0.001 |
|  | Interaction between average days addresses no-stat and COVID-19 | 0.96 | 0.19 |
|  | Phthalic anhydride | 1.02 | 0.38 |
|  | Interaction between phthalic anhydride and COVID-19 | 1.13 | <0.001 |
|  | Ethyl chloride | 1.07 | <0.001 |
|  | Interaction between ethyl chloride and COVID-19 | 1.10 | 0.001 |
|  | Nitrate | 1.10 | <0.001 |
|  | Interaction between nitrate and COVID-19 | 1.24 | <0.001 |
| Any digestive PASC | COVID-19 indicator (1=positive) | 0.61 | <0.001 |
|  | Average days addresses no-stat | 1.04 | 0.02 |
|  | Interaction between average days addresses no-stat and COVID-19 | 1.13 | <0.001 |
|  | 2,4-d, salts and esters | 1.04 | 0.07 |
|  | Interaction between COVID-19 and 2,4-d, salts and esters | 1.11 | 0.001 |
|  | Ammonium (NH4) | 1.08 | 0.01 |
|  | Interaction between NH4 and COVID-19 | 1.15 | 0.01 |
|  | COVID-19 indicator (1=positive) | 0.67 | <0.001 |
| Any general signs and symptoms PASC | Propylene oxide | 1.05 | 0.03 |
|  | Interaction between propylene oxide and COVID-19 | 1.04 | 0.24 |
|  | 2,4-d, salts and esters | 0.96 | 0.09 |
|  | Interaction between COVID-19 and 2,4-d, salts and esters | 1.18 | <0.001 |
|  | Ammonium (NH4) | 1.01 | 0.70 |
|  | Interaction between NH4 and COVID-19 | 1.05 | 0.40 |

Notes: NDI: neighborhood deprivation index.

### **eTable 6 Interactions between Contextual and Spatial Risk Factors and Individual PASC, Analysis Using OneFlorida+ Sample**

| **PASC Outcomes** | **Key Independ Variables** | **Adjusted ORs** | **P-value** |
| --- | --- | --- | --- |
| Headache | COVID-19 indicator (1=positive) | 0.82 | <0.001 |
|  | Nitrate (NO3) | 1.07 | 0.001 |
|  | Interaction between nitrate (NO3) and COVID-19 | 1.13 | <0.001 |
| Pressure ulcer of skin | COVID-19 indicator (1=positive) | 1.61 | <0.001 |
|  | Percent vacant 24 months to 36 months | 1.16 | <0.001 |
|  | Interactions between percent vacant 24 months to 36 months and COVID-19 | 1.07 | 0.28 |
| Pulmonary fibrosis | COVID-19 indicator (1=positive) | 1.49 | <0.001 |
|  | Acetamide | 1.02 | 0.42 |
|  | Interaction between acetamide and COVID-19 | 1.08 | 0.03 |
| Dyspnea | COVID-19 indicator (1=positive) | 0.92 | 0.05 |
|  | Formaldehyde | 1.01 | 0.79 |
|  | Interaction between formaldehyde and COVID-19 | 1.14 | 0.03 |
|  | 1,2-dichloropropane | 1.00 | 0.96 |
|  | Interaction between 1,2-dichloropropane and COVID-19 | 0.92 | 0.03 |
|  | Cobalt compounds | 0.99 | 0.40 |
|  | Interaction between cobalt compounds and COVID-19 | 1.07 | 0.05 |
|  | Ammonium (NH4) | 1.02 | 0.54 |
|  | Interaction between ammonium (NH4) and COVID-19 | 1.14 | 0.02 |
| Acute pharyngitis | COVID-19 indicator (1=positive) | 0.81 | <0.001 |
|  | Establishments in civic and social associations | 0.84 | <0.001 |
|  | Interaction between establishments in civic and social associations and COVID-19 | 0.99 | 0.76 |
| Chest pain | COVID-19 indicator (1=positive) | 0.85 | 0.04 |
|  | Propylene oxide | 1.20 | <0.001 |
|  | Interaction between propylene oxide and COVID-19 | 1.02 | 0.52 |
|  | Ethylene dibromide | 0.96 | 0.08 |
|  | Interaction between ethylene dibromide and COVID-19 | 0.87 | 0.003 |
|  | Mineral dust | 1.19 | 0.03 |
|  | Interaction between mineral dust and COVID-19 | 0.69 | 0.04 |
| Anemia | COVID-19 indicator (1=positive) | 0.82 | <0.001 |
|  | 2,2,4-trimethylpentane | 1.19 | <0.001 |
|  | Interaction between 2,2,4-trimethylpentane and COVID-19 | 1.01 | 0.83 |
| Edema | COVID-19 indicator (1=positive) | 0.79 | <0.001 |
|  | Average days addresses no-stat | 1.05 | 0.01 |
|  | Interaction between average days addresses no-stat and COVID-19 | 1.06 | 0.05 |
|  | Flag for low access ZCTA5 at 10 miles | 1.02 | 0.26 |
|  | Interaction between low access ZCTA5 at 10 miles and COVID-19 | 1.04 | 0.15 |
|  | Methyl chloride | 1.09 | 0.01 |
|  | Interaction between methyl chloride and COVID-19 | 1.12 | 0.06 |
|  | 2,4-d, salts and esters | 1.01 | 0.55 |
|  | Interaction between 2,4-d, salts and esters and COVID-19 | 1.09 | 0.01 |
| Abdominal pain | COVID-19 indicator (1=positive) | 0.63 | <0.001 |
|  | Average days addresses no-stat | 1.04 | 0.02 |
|  | Interaction between COVID-19 and average days addresses no-stat | 1.12 | <0.001 |
|  | 2,4-d, salts and esters | 1.13 | <0.001 |
|  | Interaction between 2,4-d, salts and esters and COVID-19 | 0.98 | 0.21 |
|  | Ammonium (NH4) | 1.04 | 0.06 |
|  | Interaction between ammonium (NH4) and COVID-19 | 1.09 | 0.008 |
| Malaise and fatigue | COVID-19 indicator (1=positive) | 0.90 | 0.07 |
|  | Low access population with housing units without vehicle access at 1 mile | 1.03 | 0.16 |
|  | Interaction between low access and COVID-19 | 1.03 | 0.32 |
|  | Phthalic anhydride | 1.06 | 0.02 |
|  | Interaction between phthalic anhydride and COVID-19 | 1.01 | 0.76 |
| Joint pain | COVID-19 indicator (1=positive) | 0.63 | <0.001 |
|  | Propylene oxide | 1.00 | 0.97 |
|  | Interaction between propylene oxide and COVID-19 | 1.20 | <0.001 |
|  | 2,4-d, salts and esters | 0.95 | 0.04 |
|  | Interaction between 2,4-d, salts and esters and COVID-19 | 1.25 | <0.001 |
|  | Ammonium (NH4) | 0.98 | 0.55 |
|  | Interaction between ammonium (NH4) and COVID-19 | 1.14 | 0.04 |
